# Latent class growth mixture modelling of HbA1C trajectories identifies individuals at high risk of developing complications of type 2 diabetes mellitus in the UK Biobank

**DOI:** 10.1101/2024.09.18.24313910

**Authors:** Dale Handley, Alexandra C Gillett, Renu Bala, Jess Tyrrell, Cathryn M Lewis

## Abstract

**Aims:** Glycated Hemoglobin A_1c_ (HbA1c) is widely used for the diagnosis and management of type 2 diabetes mellitus (T2D), with regular testing in primary care recommended every three to six months. We aimed to identify distinct, long-term HbA1c trajectories following a T2D diagnosis and investigate how these glycaemic control trajectories were associated with health-related traits and T2D complications.

**Methods:** A cohort of 12,435 unrelated individuals of European ancestry with T2D was extracted from the UK Biobank data linked to primary care records. Latent class growth mixture modelling was applied to identify classes with similar HbA1c trajectories over the 10-years following T2D diagnosis. We tested for associations of HbA1c class membership with sociodemographic factors, biomarkers, polygenic scores, and T2D-related outcomes, using logistic regression and Cox proportional hazards models.

**Results:** Six HbA1c trajectory classes were identified. The largest class (76.8%) maintained low and stable HbA1c levels over time. The other five classes demonstrated higher and more variable trajectories and included: two with parabolic shapes (starting low and distinguished by the height of their peaks), two with high initial HbA1c levels that declined over time (one rapidly, one slowly), and one class with a rapid increase in HbA1c five years after diagnosis. Younger age at T2D diagnosis, higher fasting glucose levels, higher random glucose levels, and higher body mass index polygenic score were associated with membership of these five classes. These classes were also more likely to be prescribed glucose-lowering medication at diagnosis and had fewer primary care visits in the month and year prior to diagnosis. Relative to the low and stable class, these five showed increased risks of T2D complications, including stroke (HR=1.55 [1.31-1.84]), kidney disease (HR=1.39 [1.27-1.53]), all-cause mortality (HR=1.36 [1.23-1.51]), and progression to combination therapy (HR=3.22 [3.04-3.41]) or insulin (HR=3.21 [2.89-3.55]).

**Conclusion:** Individuals with T2D who show higher and more variable HbA1c trajectories are at increased risk of developing T2D-related complications. Early identification of patients at risk, based on factors such as age at diagnosis and previous healthcare utilisation could improve patient outcomes.

## Introduction

Type 2 diabetes mellitus (T2D) is a chronic metabolic disorder characterized by persistent hyperglycemia due to insulin resistance and impaired insulin secretion (DeFranzo et al., 2015). T2D affects approximately 6.1% of the global population. This is projected to rise to 9.8% by 2050 without effective prevention (Ong *et al*., 2023).

The pathophysiological stress of T2D extends to nearly all organ systems, leading to severe complications, which are usually categorised as microvascular or macrovascular. Microvascular complications, primarily nephropathy, neuropathy, and retinopathy, result from damage to small blood vessels (Vithian *et al*., 2010). In contrast, macrovascular complications stem from damage to large vessels and are linked to accelerated atherosclerosis, leading to a high incidence of coronary artery disease, cerebrovascular disease, and peripheral artery disease among T2D patients (Einarson *et al*., 2018). Additionally, major depressive disorder (MDD) is a notable comorbidity; individuals with T2D are at greater risk of MDD compared to normoglycemic populations, and a pre-existing diagnosis of MDD increases the risk of developing T2D by 32% (Yu *et al*., 2015). Incident MDD following T2D leads to suboptimal glycaemic control, increasing the risk of vascular complications and all-cause mortality (Nefs *et* al., 2012; Khubchandani *et al*., 2023).

Optimal management of T2D, including maintaining effective glycaemic control, is important for improving health outcomes and patient quality of life (Galindo *et al*., 2023). Early and intensive glycaemic control after T2D diagnosis is essential for reducing mortality risk, while later intervention confers only modest improvements (Dailey, 2011; Adler *et al*., 2024). In clinical practice, glycaemic control is typically assessed using glycated haemoglobin (HbA1c), with measurements required every three to six months for effective monitoring. Elevated and highly variable HbA1c measures are robust predictors of complications (Pei *et al*., 2023, Lipska et al., 2023), suggesting that understanding the different patterns of HbA1c over years since T2D diagnosis can inform more personalised management strategies.

Latent class growth mixture modelling (LCGMM) is an extension of linear mixed modelling that identifies distinct subpopulations within longitudinal data which can reveal shared disease risk (Ram et al., 2009). To date, a limited number of studies have applied LCGMM to identify HbA1c trajectories over time since T2D diagnosis using routinely collected clinical data. A study by Laiteerapong *et al*. (2017) demonstrated that non-stable trajectories are associated with increased risk of microvascular complications and mortality, whilst work by Hertroijs *et al*. (2018) showed that body mass index (BMI), HbA1c and triglycerides measured at T2D diagnosis were the most important predictors of trajectory class membership. However, these studies of HbA1c trajectories focused on either the association with complications, or on identifying predictors of trajectory membership, but did not consider both comprehensively. Additionally, no existing studies have investigated associations between identified classes and healthcare utilization, MDD or genetic predictors, such as T2D and BMI polygenic scores.

Our study leverages the extensive UK Biobank (UKB) study linked to primary care records to identify subgroups of T2D by HbA1c trajectories. We applied LCGMM to identify HbA1c trajectories over a 10-year period following T2D diagnosis and identified exposures associated with class membership, considering variables measured at or prior to T2D diagnosis, including polygenic scores and MDD diagnosis. Furthermore, we examined the associations between these trajectories and T2D complications, all-cause mortality and medications, such as progression to insulin therapy. By incorporating a diverse range of class predictors and health outcomes in a large dataset, we aim to provide a comprehensive approach to understanding glycaemic control in T2D.

## Methods

### Data

The UKB is a prospective health study of ∼500,000 individuals recruited between the ages of 40 and 70 between 2006-2010 (Sudlow *et al*., 2015). UKB collects a wide range of data including physical measurements (such as HbA1c), detailed health questionnaires and genetic data. Linkage to general practice (GP) primary care data is available for 46% of UKB participants, providing diagnostic codes, clinical biochemistry tests, clinical events, and prescribing data between the years 1990 to 2017. A flowchart of study data and methods is given in Figure 1.

**Figure 1.**
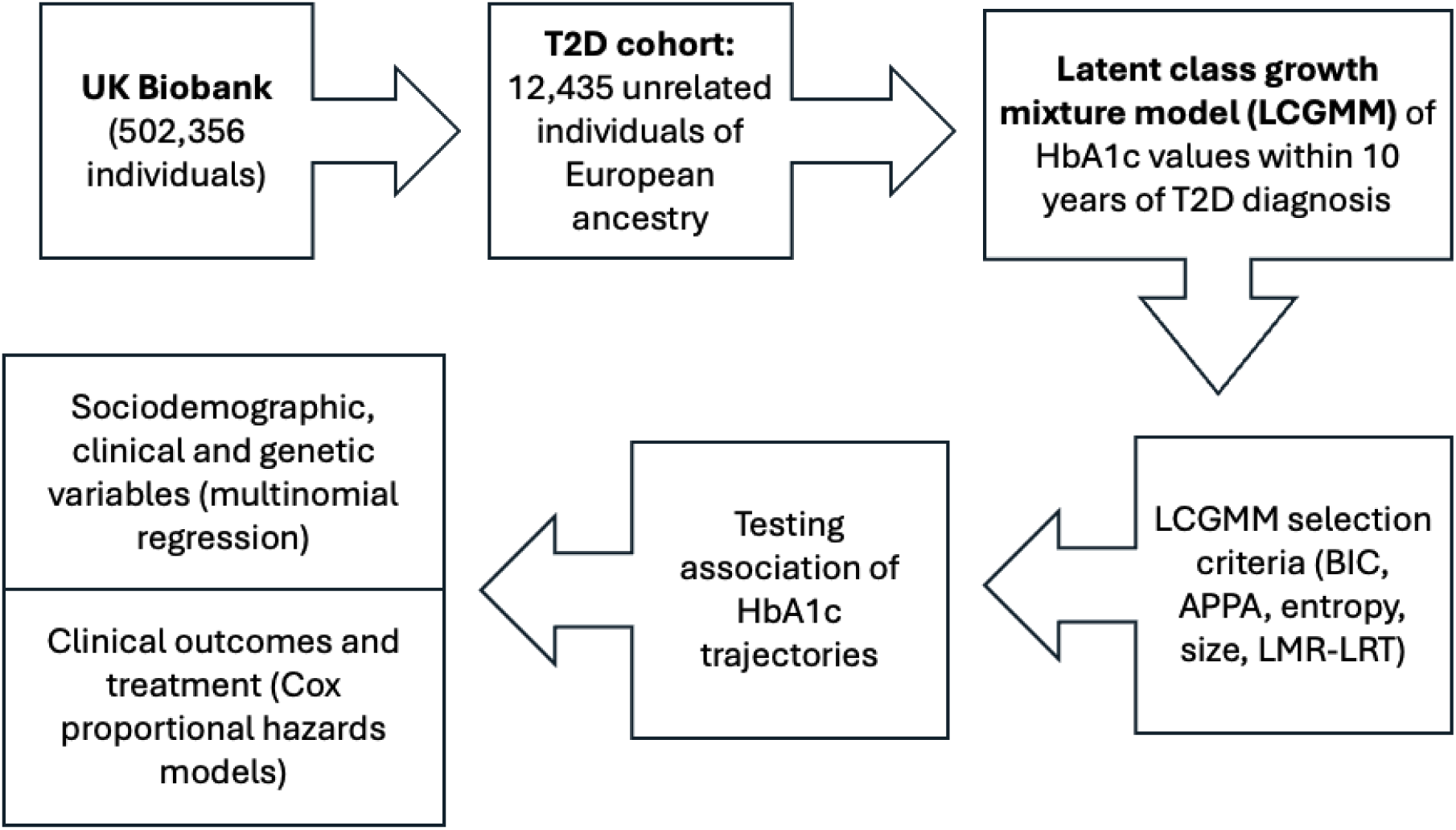
Methods flow chart. BIC, Bayesian information criteria; APPA, average posterior probability of assignment; LMR-LRT, Lo-Mendell-Rubin likelihood ratio test.

**Table 1.**
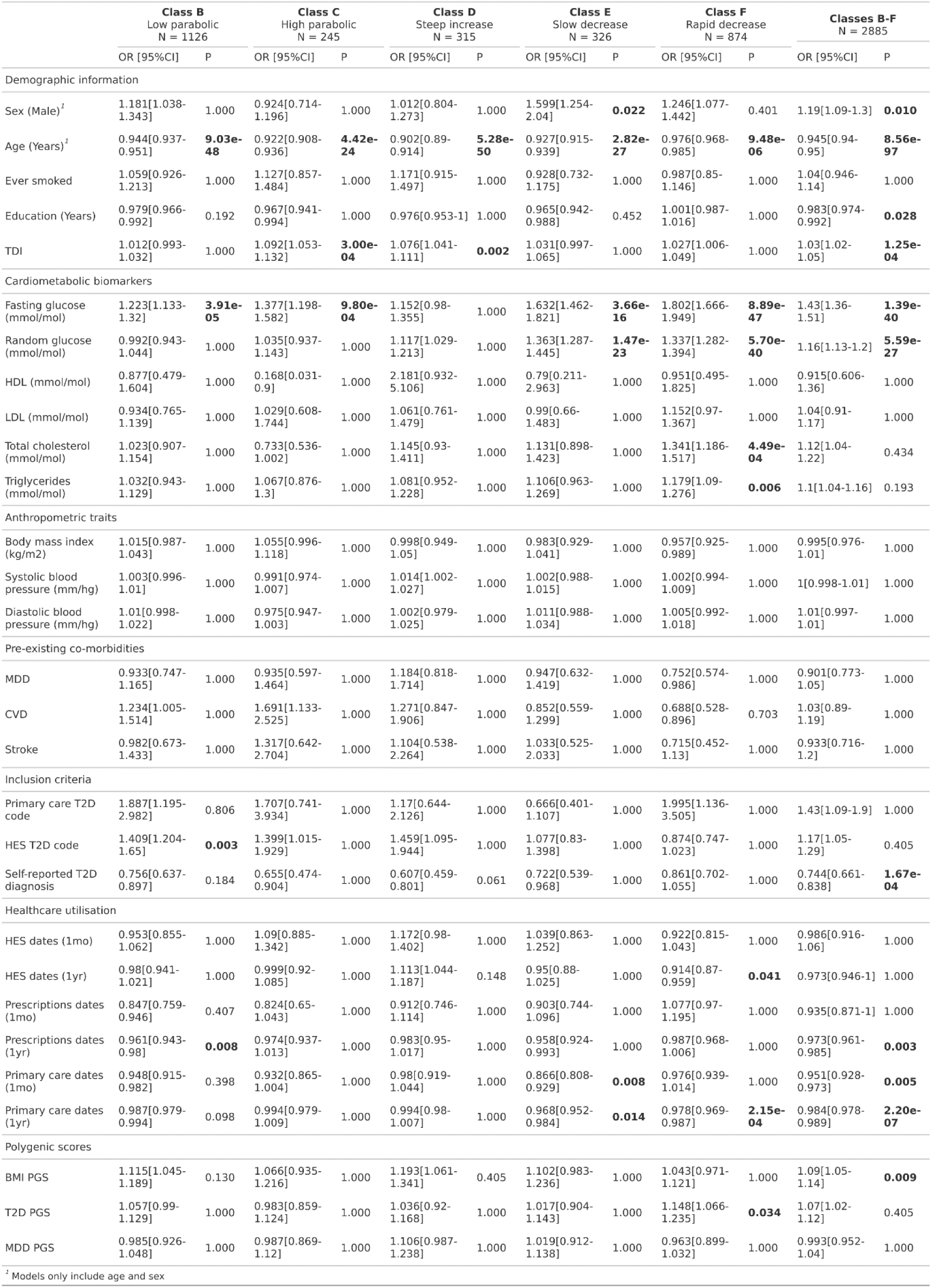
Descriptive statistics at T2D diagnosis date for all individuals, stratified by assigned class. TDI, Townsend deprivation index; HDL, high density lipoprotein; LDL, low density lipoprotein; MDD, major depressive disorder; CVD, cardiovascular disease; HES, hospital episode statistics; (1mo), only values within one month prior to diagnosis included; (1yr), only values with one year prior to diagnosis were included. IQR, inter-quartile range.

### Study population

The study population consisted of UKB participants with T2D diagnosis and linked primary care data available. Participants with T2D were identified via a previously validated approach, utilising both primary care and UKB-collected data (Gillett *et al*., 2024). Briefly, individuals must meet at least two of the following criteria: any HbA1c measurement >48 mmol/mol, any prescription for glucose-lowering medication, any self-report for T2D, a hospital episode statistics ICD9/10 code for T2D, or a GP-based T2D diagnosis. The index date was T2D diagnosis date, defined as the earliest occurrence of any of these criteria. To avoid the inclusion of mis-diagnosed non-T2D diabetes cases, we excluded individuals aged <35 years at T2D diagnosis, or those prescribed insulin within a year of diagnosis. Several additional exclusion criteria were included: 1) no recorded HbA1c measurement >38 mmol/mol within six months of diagnosis, 2) fewer than three HbA1c measurements available within 10 years of diagnosis, 3) no available genotype data, and 4) an undated MDD diagnosis.

This study utilises polygenic scores created using summary statistics from European ancestry genome-wide association studies (GWASs). Therefore, we further restricted our analyses to unrelated individuals of European ancestry (Bycroft *et al*., 2018) (Supplementary information methods S1).

### HbA1c measurements

HbA1c values were extracted from the UKB assessments and primary care records (Supplementary Table 1). All HbA1c values were converted from Diabetes Control and Complications Trial units (percentage units; DCCT) to International Federation of Clinical Chemistry and Laboratory Medicine values (mmol/mol units; IFCC). HbA1c values between 15 and 20 before transformation were removed, as it is unclear whether they represent high DCCT values or low IFCC values. As HbA1c values from the UKB assessments are systematically lower than those in the primary care records, the UKB measures were calibrated using the following equation: [calibrated HbA1c] = 0.9696[raw HbA1c] + 3.3595 (Young *et al*., 2022). Longitudinal HbA1c measurements starting from T2D diagnosis were used as the outcome when identifying subgroups of glycaemic control trajectories, with a maximum follow-up of 10 years.

### Statistical Analysis

#### Identifying HbA1C trajectories

LCGMM was used to identify classes of individuals with similar HbA1C trajectories over the first decade following their T2D diagnosis (Supplementary information methods S2). In LCGMM, convergence issues can arise when very short time intervals between observations are present. To avoid this, only one HbA1c measurement per individual every 0.1 years was included. For individuals with two measures within 0.1 years (<2% of individuals), the mean value was used. A restricted cubic spline function with three knot points was used to capture non-linear trends in HbA1c over time in the fixed effects models. Due to computational limitations, only a linear time term was included as a random effect.

To select the optimal number of classes, we started by modelling the sample as one homogeneous population and then added one class at a time. We required that a model with *k* classes must meet the following criteria to be considered for selection as the final model compared to the less complex, *k* – 1 class solution:

1. each class in the model contains a minimum of 200 individuals,
2. the Bayesian information criteria (BIC) is lower by at least 100,
3. the average posterior probability of assignment (APPA) exceeds 0.70 for all classes (Nagin, 2014),
4. the model has improved goodness of fit compared to the less complex model according to the Lo-Mendell-Rubin Adjusted Likelihood Ratio test (p < 0.05) (Lo, Mendell, and Rubin, 2001),
5. the model has a relative entropy >0.8 (Ram *et al*., 2009), and
6. the model has a mismatch value close to zero for all classes.

Class characteristics at index date were summarised (median and interquartile range; counts and proportions), with overall p-values for between group differences assessed using one-way ANOVA for continuous characteristics and likelihood ratio tests for dichotomous characteristics.

#### Testing for association of exposures with HbA1c trajectories

Two complementary approaches were used to examine the associations between exposures, measured at or prior to T2D diagnosis, and class membership. Firstly, we applied multinomial logistic regression (MLR) to identify exposures associated with each class relative to the reference class (the most common class). Secondly, we utilised logistic regression to the most common class (the reference) versus all other identified classes combined. This allowed us to identify exposures associated with deviating from the most common HbA1c trajectory. All models were adjusted for genetic sex and age at T2D diagnosis. Holm-Bonferroni correction was applied to account for multiple testing, with adjusted p-values presented.

##### Exposures

A range of demographic, biomarker, pre-existing diagnoses, polygenic scores, and healthcare utilisation variables were considered. Demographic exposures considered were genetic sex, number of education years, smoking status (ever vs. never) and Townsend deprivation index (TDI), as defined by UKB fields (Supplementary Table 1). Age at T2D diagnosis was defined as the difference between a participant’s T2D diagnosis date and date of birth. As the day of birth is unavailable in the UKB, this was set to the 1^st^ of the month for all participants. The following biomarker variables were included, created following definitions provided by Kim *et al*., 2023 and using primary care and UKB assessment data: BMI, fasting blood glucose, random blood glucose, diastolic blood pressure (DBP), systolic blood pressure (SBP), total blood cholesterol, high density lipoprotein, low density lipoprotein, and triglycerides. For all biomarkers, only measurements taken up to one-month before T2D diagnosis were considered. Healthcare utilisation exposures were the number of GP visits, the number of unique hospital episode statistics (HES) dates and the number of unique prescription dates. The number of GP visits was calculated using the number of unique dates for which an individual had at least one primary care code for both one month and one year prior to T2D diagnosis. The total number of unique Hospital dates, and total number of unique prescription dates were calculated similarly. Pre-existing diagnoses considered were stroke, cardiovascular disease (CVD), and MDD, defined using previously described methods based solely on primary care records (Fabbri *et al*., 2021; Kim *et al*., 2023). Polygenic scores for T2D, BMI, and MDD were generated and validated in UKB using PRS-CS via the GenoPred pipeline (Supplementary Information: methods S3, S4, and results S1) (Ge *et* al., 2019; Pain *et al*., 2024).

#### Testing for association of HbA1c trajectories with secondary outcomes

Time-to-event analysis was performed using Cox proportional hazards modelling to determine whether class membership increased the risk of developing secondary outcomes. The time of entry was defined as the T2D diagnosis date. When the secondary outcome was all-cause mortality, the censoring date was the last date the national death registry linkage was updated (22/11/2021). For all other outcomes, the censoring date was the earlier of the date of death or the last date of data collection. All models were adjusted for age at T2D diagnosis, genetic sex, years of education, TDI, and ever-smoked status. The reference group for class membership was the modal class. All p-values are reported as Holm-Bonferroni corrected values.

##### Secondary outcomes

The following eight phenotypes were used as outcomes in the time-to-event analyses: all-cause mortality, all cardiovascular disease combined, diabetic kidney disease, all strokes combined, peripheral artery disease, MDD, progression to combination T2D therapy, and progression to insulin. All-cause mortality was defined using the linked Office for National Statistics (ONS) death registry. Peripheral artery disease was defined according to Klarin *et al*., 2019 and MDD according to *Fabbri et al*., 2021. All other outcomes were obtained using previously defined UKB inclusion and exclusion code lists and fields for T2D complications (Kim *et al*., 2023).

Two medication phenotypes were included as measures of T2D disease progression rate (Fonseca *et al*., 2009). Firstly, “Progression to combination therapy” was defined as the time between T2D diagnosis and the first prescription record for a non-metformin medication and calculated for individuals who had at least one prescription record for metformin. Secondly, “Progression to insulin” was calculated for all individuals as the time from T2D diagnosis to the first prescription of insulin or an insulin-containing medication. Full code lists for T2D medications are available from Gillett *et al*., 2024.

#### Software and code availability

All analyses were performed using R v4.2.0. LCGMM and model selection utilised the *lcmm* and *LCTMToolkit* packages (Proust-lima *et al*., 2017; Lennon *et al*., 2018). MLR was conducted using the *nnet* package (Ripley *et al*., 2023). The *survival* package was used for Cox proportional hazards modelling (Therneau *et al*., 2000). All other figures and tables were created using *ggplot2* (Wickham H, 2016) and *gt* (Lannone *et al*., 2024). Code for this project is available at https://github.com/dale-handley/UKBIOBANK-HbA1c-LCGMM.

## Results

### Cohort characteristics

In total, 12,435 individuals met the study inclusion criteria (Supplementary Figure 1). The cohort was predominantly male (61.6%), with an average age at T2D diagnosis of 59.7 years. The median follow-up time was eight years, with a median of 13 HbA1c measurements available.

### Latent class growth mixture modelling

In the analysis of 10-year HbA1c trajectories among individuals newly diagnosed with T2D, six models converged within the designated timeframe. The model with six classes was the best fitting, meeting the predefined selection criteria (Supplementary Tables 2 and 3). Most individuals were assigned to Class A (76.8%), which had the lowest initial HbA1c of any class, which remained low and slowly increased over time (‘low and stable’). This class is the reference class in subsequent analyses. The other five classes had distinct trajectories that were typically higher and more variable: (1) Class B (9.06%) showed low HbA1c levels at diagnosis that moderately increased until year five, then gradually returned to initial levels (‘low parabolic’), (2) Class C (1.97%) started low, rapidly increased until 4.5 years after diagnosis (‘high parabolic’), (3) Class D (2.53%) had a steep increase in HbA1c after two years (‘steep increase’), (4) Class E (2.62%) started high and slowly decreased during the 10 years following T2D diagnosis (‘slow decrease’) and (5) Class F (7.03%) had high HbA1c levels at diagnosis that rapidly decreased over the first 18 months, then gradually increased from the fourth year (‘rapid decrease’). See Figure 2 and Table 2 for details.

**Figure 2.**
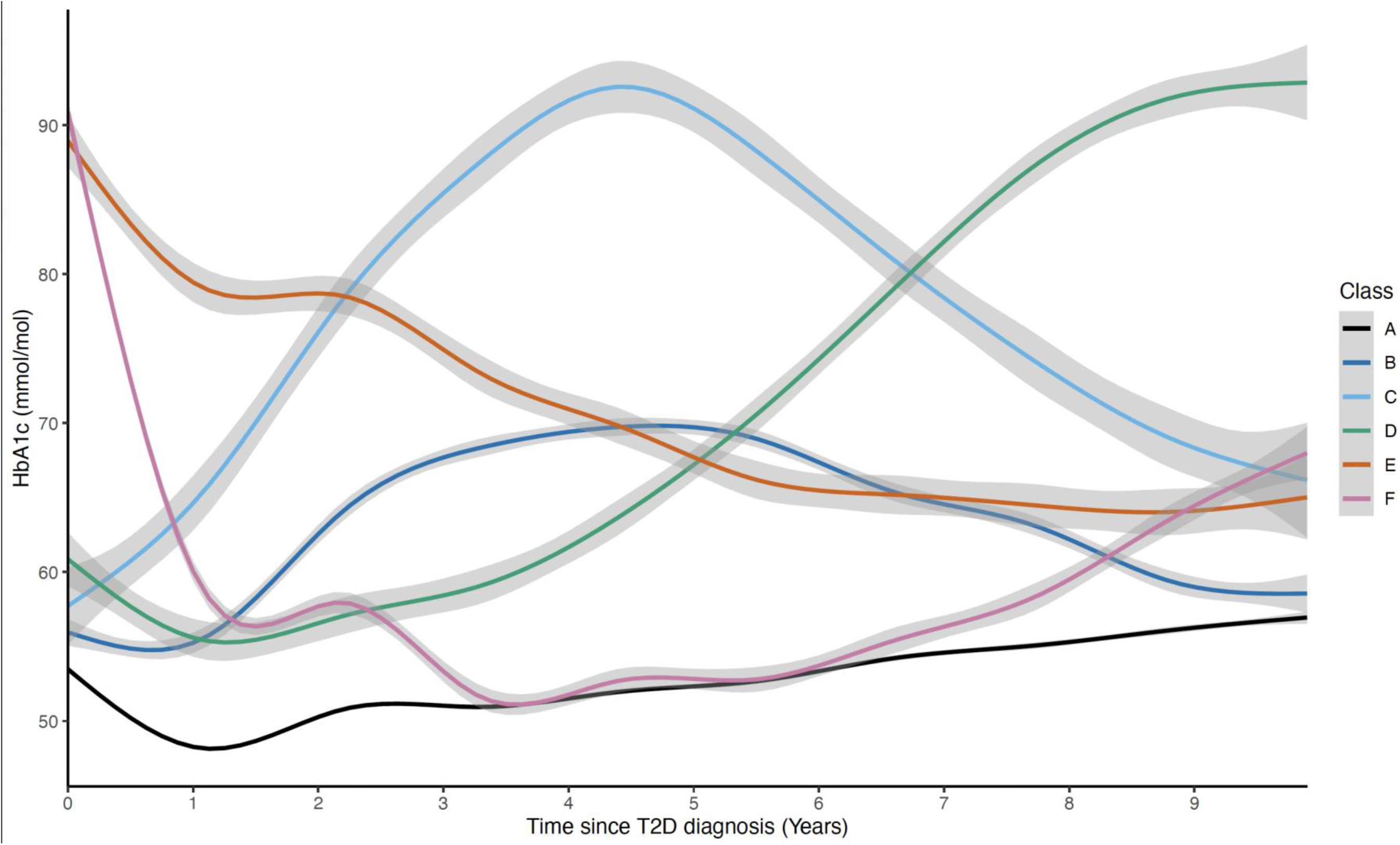
Mean HbA1c trajectories during the first 10-years post-T2D diagnosis for all classes. Shaded area represents 95% confidence intervals.

**Table 2:**
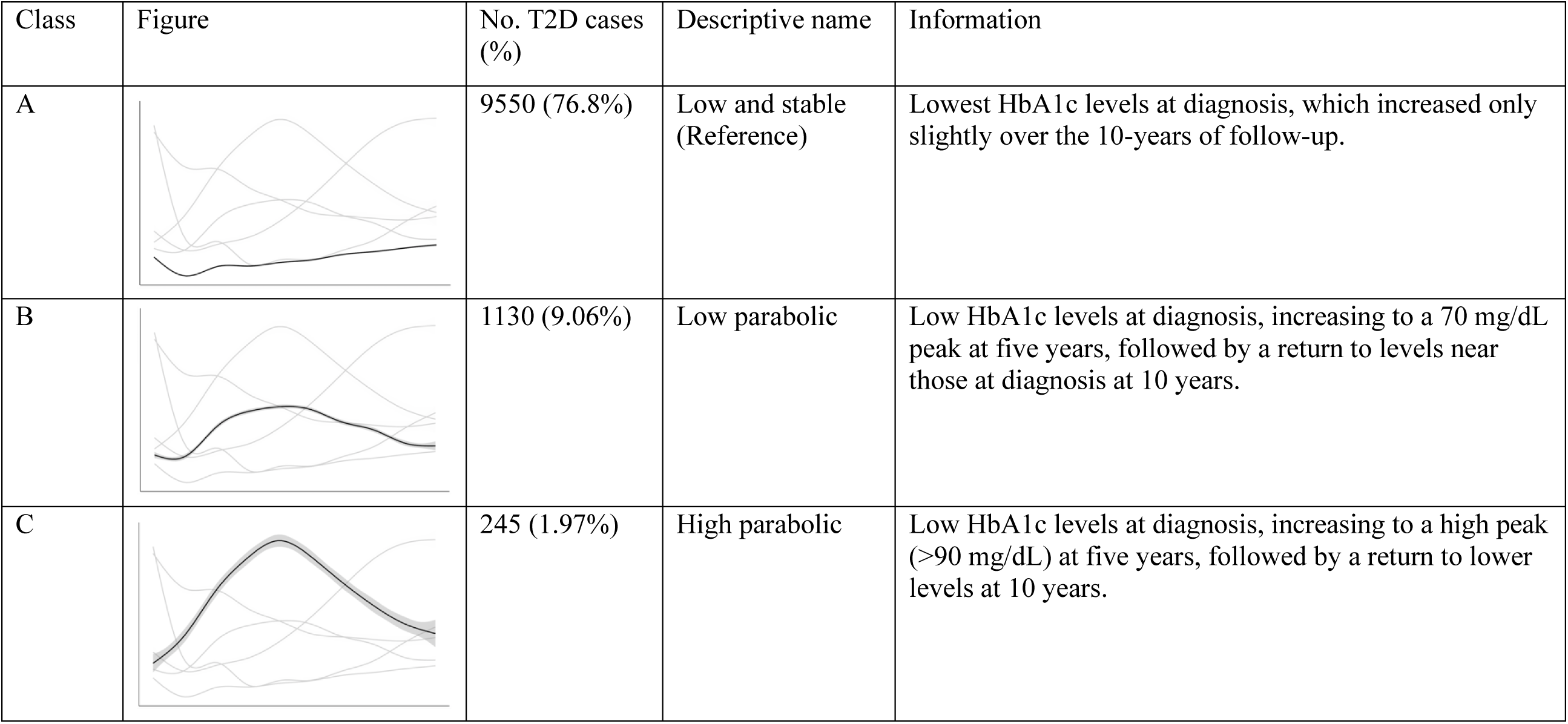
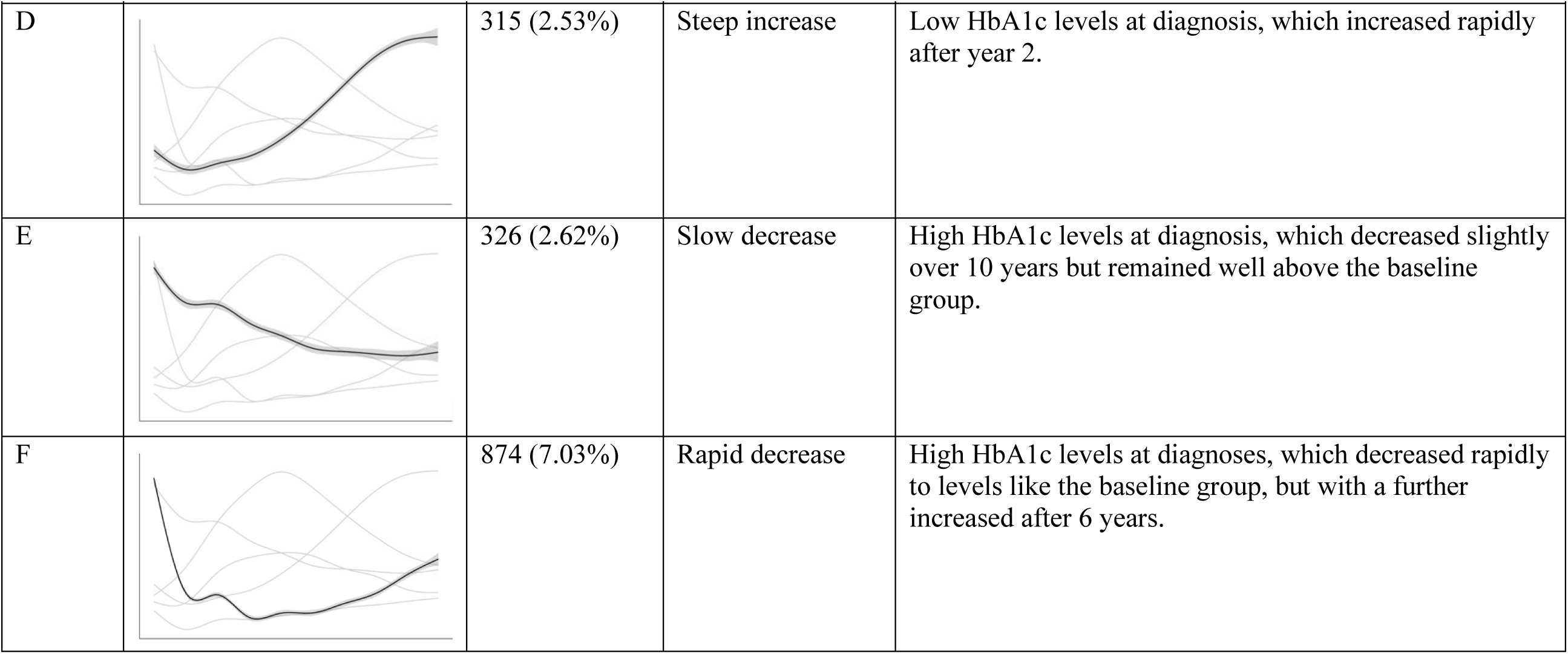
Descriptives of the six-classes in the latent class growth mixture model, ordered by the predicted HbA1c level at diagnosis.

| Class | Figure | No. T2D cases (%) | Descriptive name | Information |
| --- | --- | --- | --- | --- |
| A |  | 9550 (76.8%) | Low and stable (Reference) | Lowest HbA1c levels at diagnosis, which increased only slightly over the 10-years of follow-up. |
| B |  | 1130 (9.06%) | Low parabolic | Low HbA1c levels at diagnosis, increasing to a 70 mg/dL peak at five years, followed by a return to levels near those at diagnosis at 10 years. |
| C |  | 245 (1.97%) | High parabolic | Low HbA1c levels at diagnosis, increasing to a high peak (>90 mg/dL) at five years, followed by a return to lower levels at 10 years. |
| D |  | 315 (2.53%) | Steep increase | Low HbA1c levels at diagnosis, which increased rapidly after year 2. |
| E |  | 326 (2.62%) | Slow decrease | High HbA1c levels at diagnosis, which decreased slightly over 10 years but remained well above the baseline group. |
| F |  | 874 (7.03%) | Rapid decrease | High HbA1c levels at diagnoses, which decreased rapidly to levels like the baseline group, but with a further increase after 6 years. |

### Exposures associated with class membership

Compared to Class A (‘low and stable’), membership of all other classes (B-F) was associated with a younger age at T2D diagnosis (Table 3). Additionally, higher fasting glucose levels at T2D diagnosis were associated with increased odds of class membership relative to Class A, in all classes except Class D (‘steep increase’). Higher Townsend deprivation index was linked with increased odds of belonging to Classes C (‘high parabolic’; OR=1.09, 95% confidence interval [1.05,1.13]) and D (OR=1.10 [1.04-1.11]), whilst higher random glucose levels at T2D diagnosis increased the odds of membership to Class E (‘slow decrease’; OR=1.36 [1.29-1.45]) and Class F (‘rapid decrease’; OR=1.34 [1.28-1.39]) relative to Class A. Males were more likely than females to belong to Class E compared to Class A (OR=1.60 [1.25-2.04]). Finally, membership of Class F was associated with higher levels of total cholesterol (OR=1.34 [1.19-1.52]), triglycerides (OR=1.18 [1.09-1.28]) and T2D polygenic score (OR=1.15 [1.07-1.24]).

**Table 3.**
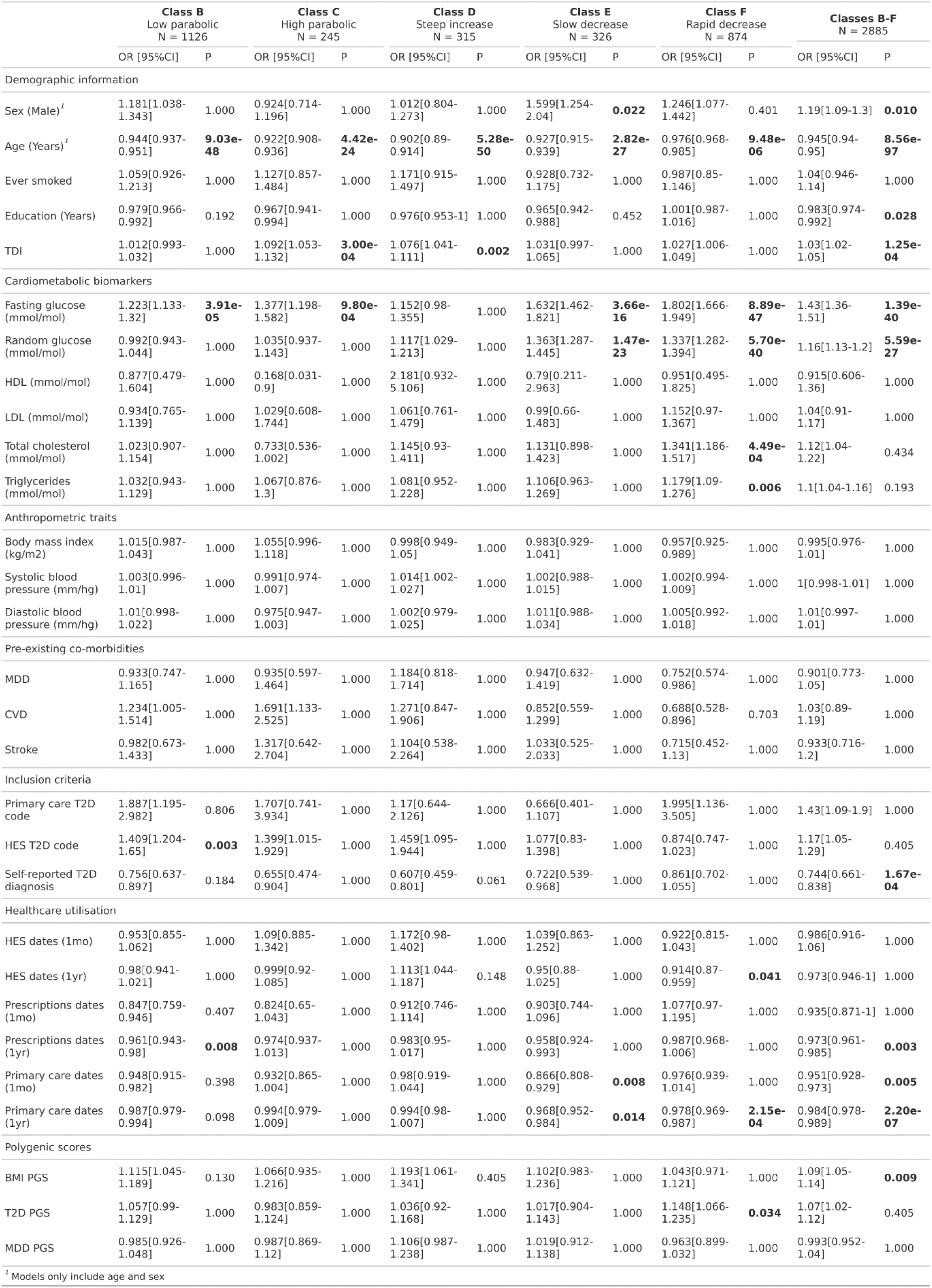
Associations between exposures and HbA1c class membership, adjusted for sex and age at T2D diagnosis. The effect size is reported as odds ratio and 95% confidence intervals in square brackets for each class, compared to the reference class (A, ‘low and stable’). To generate the Overall non-reference class models (B-F), all individuals who were not assigned to class A were considered a single class. HDL, high density lipoprotein; LDL, low density lipoprotein; BMI, Body mass index; MDD, Major depressive disorder; CVD, cardiovascular disease. Values in bold represent Holm-Bonferroni adjusted p-values which are statistically significant (P < 0.05 after testing correction applied).

Comparing individuals in reference Class A with all other classes combined showed that membership of classes B-F was associated with a younger age at T2D diagnosis (OR=0.95 [0.94-0.95]), being male (OR=1.19 [1.09-1.30]), having fewer years in education (OR=0.98 [0.97-0.99]) and higher levels of TDI (OR=1.03 [1.02-1.05]), fasting glucose (OR=1.43 [1.36-1.51]), random glucose (OR=1.16 [1.13-1.20]) and BMI polygenic score (OR=1.09 [1.05-1.14]).

### Source of T2D diagnosis criteria and healthcare utilisation is associated with class membership

Given the high HbA1c intercept values observed in Classes E and F, we examined the association between class membership and healthcare utilisation, as well as the source of T2D diagnosis, to assess whether diagnosis in some classes may have occurred secondary to another clinical investigation or during emergency care (Tables 2 and 3). Compared to Class A, individuals in the non-reference class were less likely to self-report a diagnosis of T2D (OR=0.74 [0.66 – 0.84]). Fewer hospital episode dates in the year prior to T2D diagnosis were also associated with membership of Class F (OR=0.91 [0.87-0.96]). Additionally, having fewer primary care dates within the year before diagnosis was associated with higher odds of belonging to Class E (OR=0.97 [0.95-0.98]) and Class F (OR=0.98 [0.97-0.99]) compared to Class A. Additionally, belonging to a non-reference class was associated with fewer GP visits in both the year (OR=0.98 [0.98-0.99]) and month (OR=0.95 [0.93-0.97]) prior to T2D diagnosis, and with fewer prescriptions in the year before diagnosis (OR=0.97 [0.96-0.99]), indicating that reduced healthcare utilisation is associated with non-reference class membership.

### Class membership associated with secondary outcomes

Secondary outcomes were strongly associated with class membership after correcting for multiple testing (Figure 4; supplementary tables 4 and 5). All non-reference classes showed an increased risk of developing diabetic retinopathy relative to Class A. Class E (‘slow decrease’) had the highest risk, b1.43 (95% CI: 1.26-1.61) condition compared to those in Class A, whilst Class B (‘low parabolic’) had the lowest relative risk (HR=1.25 [1.13-1.38]). Higher rates of all-cause mortality, stroke and CVD events were seen in classes C (‘high parabolic’), D (‘steep increase’) and E compared to Class A. For example, for stroke, the hazard rate ranged from 2.07 (95% CI:1.28-3.34) for Class C, to 2.72 (95% CI: 1.88-3.94) for Class D, compared to class A. All classes except Class E were at increased risk of developing kidney disease compared to Class A.

**Figure 4.**
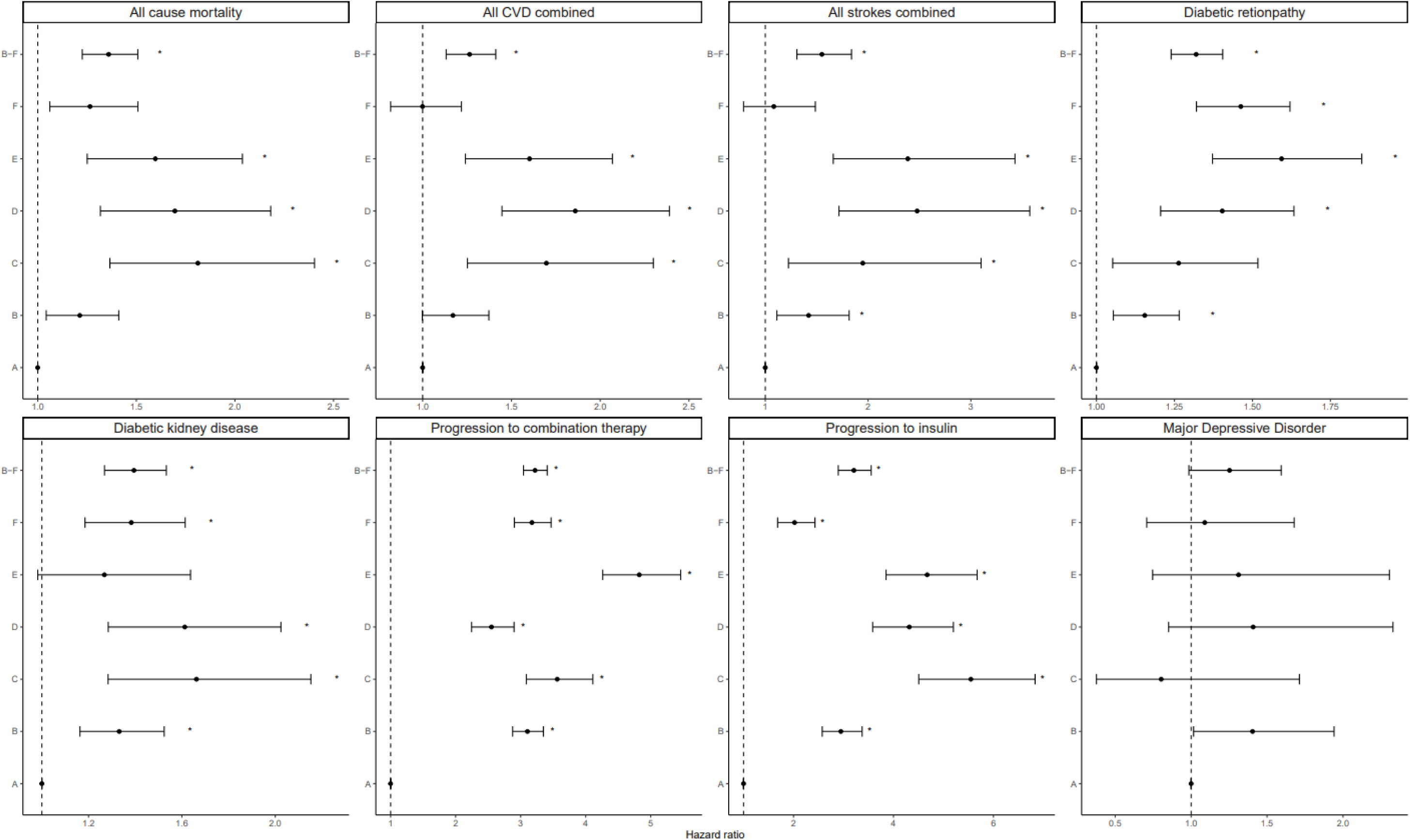
Association of class membership with T2D outcomes, using time-to-event analysis. Error bars represent 95% confidence intervals. asterisks represent statistically significant p-values after applying Bonferroni-Holm correction (P < 0.05). CVD, cardiovascular disease. To generate the Overall non-reference class models (B-F), all individuals who were not assigned to class A were considered a single class.

All classes were associated with progression to dual therapy and to insulin. Class F (‘rapid decrease’) had the lowest estimated HR for both medication outcomes, being 1.77 (95% CI: 1.45–2.16) times more likely to progress to combination therapy and 1.87 (95% CI: 1.63–2.14) times more likely to progress to insulin compared to Class A. Class E had the highest risk of progressing to combination therapy (HR=4.04 [3.38–4.81]) and Class C had the highest risk of progressing to insulin (HR=5.67 [4.59– 7.02]). There was no difference in the risk of MDD diagnosis for any class compared to the reference.

When Classes B-F were combined into one group (atypical trajectories) and compared to Class A (typical), we observed that belonging to an atypical trajectory (Classes B-F) was associated with an increased risk of experiencing all secondary outcomes considered, except for MDD.

## Discussion

In this study, we utilised the extensive data available in the UKB and identified six classes of individuals of European ancestry with T2D based on their HbA1c trajectories over a mean follow-up of 8.1 years from diagnosis. The reference class (A; ‘low and stable’) represents a trajectory previously observed in other studies. This class has the lowest initial HbA1c levels of all classes and experiences a dip around one year after diagnosis, before rebounding and slowly increasing thereafter (Hertroijs *et al* 2018; Gillett *et al*., 2024). Classes B (‘low parabolic’) and C (high parabolic’) represent individuals who experience increases in HbA1c during the first five years post-diagnosis, followed by a decline, with Class C experiencing a more pronounced increase and subsequent decline than Class B. Classes E (‘slow decrease’) and F (‘rapid decrease’) contain individuals with high HbA1c levels at diagnosis, that decrease over time, with class F having a higher initial HbA1c and a steeper decline. Class D (‘steep increase’) have a moderately high initial HbA1c, with a rapid increase approximately four years post-diagnosis. Overall, 23.2% of participants were members of Classes B-F (non-reference/ atypical classes), characterised by higher and more variable HbA1c trajectories compared to the low and stable trend observed in Class A (reference/ typical class).

Younger age of T2D onset and higher fasting blood glucose at T2D diagnosis were associated with class membership for nearly all classes, compared to Class A. While the association with fasting glucose is expected due to its strong correlation with HbA1c, the link between younger age of T2D onset and class membership indicates that individuals who develop T2D at a younger age tend to have poorer glycaemic control and a higher risk of developing complications. Possible explanations for this include having higher adiposity, showing lower adherence to T2D medications, and/ or having a genetically distinct phenotype which is associated with different risks of T2D complications (Noordam *et al*., 2021, Shahabi *et* al., 2023). This finding aligns with previous studies, which have identified younger age of T2D onset as a risk factor for developing vascular complications (Wilmot *and* Idris, 2014).

In the all-classes combined analysis, higher BMI polygenic score was associated with non-reference class membership, but phenotypically measured BMI was not. This could suggest a pleiotropic genetic effect of the BMI polygenic score on body fat composition, which is strongly associated with the development of T2D and complications (Li *et al*., 2021; Xu *et al*., 2024; Guo *et al*., 2024). Additionally, the BMI polygenic score may be capturing changes in BMI that occur alongside the HbA1c trajectories which may not be captured by a single phenotypic BMI measurement (Khera *et al*., 2019).

In the healthcare utilization analysis, we found a strong link between belonging to a non-reference class and reduced GP visits before T2D diagnosis. Since increased GP visitation is associated with lower hospitalization rates in T2D patients and early diagnosis is crucial for glycaemia management, these results suggest that lower healthcare utilization prior to diagnosis could lead to a more advanced T2D at diagnosis, poorer glycaemic control, and higher risk of post-diagnosis complications (Ha *et al*., 2020).

The time-to-event analysis of secondary outcomes showed that all non-standard classes were at increased risk of developing all disease outcomes, except MDD, compared to Class A. While the association between atypical HbA1c trajectories and risk of T2D complications is well established, we identified three classes (C, D, E) at notably elevated risk of diabetes-related complications and all-cause mortality. The observed differences between Classes E and F (slow vs. rapid decrease) demonstrates total exposure to high HbA1c levels and early glycaemic management as important risk factors for the development of complications, as reported previously (Boye *et al*., 2022). Similarly, the increased HbA1c variability when comparing Classes B and C (low vs. high parabolic) underscores the need for longer term and more intensive HbA1c management upon the observation of rapidly increasing HbA1c. Class D may represent failure to respond to second line T2D therapies or therapeutic inertia, where failure to intensify or de-intensify glycaemic therapies with adequate timing leads to poorer long-term glycaemic control and increased risk of complications (Rodriguez *et al*., 2024).

This UKB study of T2D HbA1c trajectories offers several advantages over previous research. First, the strict T2D definition increases the likelihood of including only true T2D cases. Second, the large sample sizes, stringent model selection, and flexible time modelling ensure that LCGMM-generated trajectories represent meaningful HbA1c subclasses (Mesidor et al., 2022). Third, we consider a broad range of exposures and secondary outcomes, allowing a comprehensive investigation of how these trajectories develop and affect the clinical course of T2D in the UKB. Finally, this is the first study to incorporate genetic information into LCGMM, revealing a novel association between the BMI polygenic score, but not BMI, and atypical HbA1c trajectories, as well as a class-specific association between the T2D polygenic score and Class F.

However, our study has several limitations. First, LCGMM is computationally intensive. It is therefore possible that with more computational power, a model with more classes could have been selected. However, this is unlikely as the improvements in goodness of fit and class stability by including more classes was small after inclusion of a sixth class (Supplementary Table 3). Second, there is a healthy volunteer bias in the UKB, meaning these individuals may not be truly representative of the UK-based T2D population. A higher proportion of T2D patients may therefore be assigned to non-reference classes in a more representative UK sample. Cohorts that do not rely on individual-level recruitment, such as the Clinical Practice Research Datalink, should be used to validate our results. Third, T2D diagnosis criteria have changed substantially over the past two decades, with increased population-wide T2D screening and monitoring. This could introduce cohort-specific effects related to the start date of medical records in the UKB. Fourth, despite the large number of participants in our study, only 8% and 4% had prevalent and incident major depressive disorder (MDD) respectively, limiting our ability to fully determine the relationship between HbA1c class trajectories and MDD. Finally, due to limited sample size for individuals of non-European ancestry, and the lack of availability for well-powered genetic studies in individuals in this group, we excluded individuals who were non-European. As there is known heterogeneity by ancestry for T2D disease progression and outcomes (Spanakis *et al*., 2013; Vasishta *et al*., 2022), further studies should focus on extending these methods to individuals of non-European ancestry.

In conclusion, six classes with distinct HbA1c trajectories were identified in 12,435 individuals of European ancestry with T2D, using the UKB linked to primary care data. Participants in non-reference classes (23%) had increased risk of developing T2D complications, including all-cause mortality. Younger age at T2D diagnosis and lower healthcare utilisation prior to diagnosis were identified as important risk factors for following an atypical trajectory. This suggests that the risk of developing complications is related to lower healthcare utilisation prior to T2D diagnosis. Additionally, although all individuals in this study that followed an atypical trajectory were at higher risk of developing complications, we identify ∼7% of participants (Classes C, D, E) who displayed highly accelerated T2D progression, as defined by a greater degree of medication transition and a higher risk of developing T2D complications and all-cause mortality. In summary, this study shows that participants who diverge from the typical T2D HbA1c trajectory can be identified as targets for intensive intervention.

## Data Availability

Data used in this project is available via application to the UK Biobank.

## Acknowledgements

This study was supported by the National Institute for Health and Care Research Exeter Biomedical Research Centre and by the NIHR Maudsley Biomedical Research Centre at South London and Maudsley NHS Foundation Trust and King’s College London. The views expressed are those of the author(s) and not necessarily those of the NIHR or the Department of Health and Social Care. This research has been conducted using the UK Biobank resource under application number 82087.

## Notes

### Competing Interest Statement

The authors have declared no competing interest.

### Funding Statement

This paper represents independent research part-funded by the National Institute for Health Research (NIHR) Maudsley Biomedical Research Centre at South London and Maudsley NHS Foundation Trust and King's College London. The views expressed are those of the author(s) and not necessarily those of the NHS, the NIHR, or the Department of Health and Social Care. The authors acknowledge use of the research computing facility at King's College London, CREATE. This research as supported by the Medical Research Council (MR/S0151132; MR/N015746). JT is supported by an Academy of Medical Sciences (AMS) Springboard award, which is supported by the AMS, the Wellcome Trust, GCRF, the Government Department of Business, Energy and Industrial strategy, the British Heart Foundation and Diabetes UK [SBF004\1079].

## References

Adler, A.I., Coleman, R.L., Leal, J., Whiteley, W.N., Clarke, P., Holman, R.R., 2024. Post-trial monitoring of a randomised controlled trial of intensive glycaemic control in type 2 diabetes extended from 10 years to 24 years (UKPDS 91). The Lancet 404, 145–155. 10.1016/S0140-6736(24)00537-3

An, X., Zhang, Y., Sun, W., Kang, X., Ji, H., Sun, Y., Jiang, L., Zhao, X., Gao, Q., Lian, F., Tong, X., 2024. Early effective intervention can significantly reduce all-cause mortality in prediabetic patients: a systematic review and meta-analysis based on high-quality clinical studies. Front Endocrinol (Lausanne) 15, 1294819. 10.3389/fendo.2024.1294819

Boye, K.S., Thieu, V.T., Lage, M.J., Miller, H., Paczkowski, R., 2022. The Association Between Sustained HbA1c Control and Long-Term Complications Among Individuals with Type 2 Diabetes: A Retrospective Study. Adv Ther 39, 2208–2221. 10.1007/s12325-022-02106-4

Bycroft, C., Freeman, C., Petkova, D., Band, G., Elliott, L.T., Sharp, K., Motyer, A., Vukcevic, D., Delaneau, O., O’Connell, J., Cortes, A., Welsh, S., Young, A., Effingham, M., McVean, G., Leslie, S., Allen, N., Donnelly, P., Marchini, J., 2018. The UK Biobank resource with deep phenotyping and genomic data. Nature 562, 203–209. 10.1038/s41586-018-0579-z

Ceriello, A., Prattichizzo, F., Phillip, M., Hirsch, I.B., Mathieu, C., Battelino, T., 2022. Glycaemic management in diabetes: old and new approaches. Lancet Diabetes Endocrinol 10, 75–84. 10.1016/S2213-8587(21)00245-X

Choe, E.K., Shivakumar, M., Lee, S.M., Verma, A., Kim, D., 2022. Dissecting the clinical relevance of polygenic risk score for obesity – a cross-sectional, longitudinal analysis. Int J Obes (Lond) 46, 1686–1693. 10.1038/s41366-022-01168-2

Dailey, G., 2011. Early and intensive therapy for management of hyperglycemia and cardiovascular risk factors in patients with type 2 diabetes. Clin Ther 33, 665–678. 10.1016/j.clinthera.2011.04.025

Davies, M.J., Aroda, V.R., Collins, B.S., Gabbay, R.A., Green, J., Maruthur, N.M., Rosas, S.E., Prato, S.D., Mathieu, C., Mingrone, G., Rossing, P., Tankova, T., Tsapas, A., Buse, J.B., 2022. Management of Hyperglycemia in Type 2 Diabetes, 2022. A Consensus Report by the American Diabetes Association (ADA) and the European Association for the Study of Diabetes (EASD). Diabetes Care 45, 2753. 10.2337/dci22-0034

DeFronzo, R.A., Ferrannini, E., Groop, L., Henry, R.R., Herman, W.H., Holst, J.J., Hu, F.B., Kahn, C.R., Raz, I., Shulman, G.I., Simonson, D.C., Testa, M.A., Weiss, R., 2015. Type 2 diabetes mellitus. Nat Rev Dis Primers 1, 15019. 10.1038/nrdp.2015.19

Ds, N., Bl, J., Vl, P., Re, T., 2018. Group-based multi-trajectory modeling. Statistical methods in medical research 27. 10.1177/0962280216673085

Einarson, T.R., Acs, A., Ludwig, C., Panton, U.H., 2018. Prevalence of cardiovascular disease in type 2 diabetes: a systematic literature review of scientific evidence from across the world in 2007–2017. Cardiovascular Diabetology 17, 83. 10.1186/s12933-018-0728-6

Fabbri, C., Hagenaars, S.P., John, C., Williams, A.T., Shrine, N., Moles, L., Hanscombe, K.B., Serretti, A., Shepherd, D.J., Free, R.C., Wain, L.V., Tobin, M.D., Lewis, C.M., 2021. Genetic and clinical characteristics of treatment-resistant depression using primary care records in two UK cohorts. Molecular Psychiatry 26, 3363. 10.1038/s41380-021-01062-9

Fonseca, V.A., 2009. Defining and characterizing the progression of type 2 diabetes. Diabetes Care 32 Suppl 2, S151–156. 10.2337/dc09-S301

Galindo, R.J., Trujillo, J.M., Low Wang, C.C., McCoy, R.G., 2023. Advances in the management of type 2 diabetes in adults. BMJ Med 2, e000372. 10.1136/bmjmed-2022-000372

Ge, T., Chen, C.-Y., Ni, Y., Feng, Y.-C.A., Smoller, J.W., 2019. Polygenic prediction via Bayesian regression and continuous shrinkage priors. Nat Commun 10, 1776. 10.1038/s41467-019-09718-5

Gillett, A.C., Hagenaars, S.P., Handley, D., Casanova, F., Young, K.G., Green, H., Lewis, C.M., Tyrrell, J., 2024. The impact of major depressive disorder on glycaemic control in type 2 diabetes: a longitudinal cohort study using UK Biobank primary care records. BMC Medicine 22, 211. 10.1186/s12916-024-03425-9

Guo, N., Shi, H., Zhao, H., Abuduani, Y., Chen, D., Chen, X., Wang, H., Li, P., 2024. Causal relationships of lifestyle behaviours and body fat distribution on diabetic microvascular complications: a Mendelian randomization study. Front. Genet. 15. 10.3389/fgene.2024.1381322

Ha, N.T., Harris, M., Preen, D., Moorin, R., 2020. Original research: Time protective effect of contact with a general practitioner and its association with diabetes-related hospitalisations: a cohort study using the 45 and Up Study data in Australia. BMJ Open 10. 10.1136/bmjopen-2019-032790

Hertroijs, D.F.L., Elissen, A.M.J., Brouwers, M.C.G.J., Schaper, N.C., Köhler, S., Popa, M.C., Asteriadis, S., Hendriks, S.H., Bilo, H.J., Ruwaard, D., 2018. A risk score including body mass index, glycated haemoglobin and triglycerides predicts future glycaemic control in people with type 2 diabetes. Diabetes, Obesity & Metabolism 20, 681. 10.1111/dom.13148

Iannone, R., Cheng, J., Schloerke, B., Hughes, E., Lauer, A., Seo, J., Brevoort, K., Roy, O., 2020. gt: Easily Create Presentation-Ready Display Tables. 10.32614/CRAN.package.gt

Khera, A.V., Chaffin, M., Wade, K.H., Zahid, S., Brancale, J., Xia, R., Distefano, M., Senol-Cosar, O., Haas, M.E., Bick, A., Aragam, K.G., Lander, E.S., Smith, G.D., Mason-Suares, H., Fornage, M., Lebo, M., Timpson, N.J., Kaplan, L.M., Kathiresan, S., 2019. Polygenic prediction of weight and obesity trajectories from birth to adulthood. Cell 177, 587–596.e9. 10.1016/j.cell.2019.03.028

Khubchandani, J., Banerjee, S., Gonzales-Lagos, R., Szirony, G.M., 2023. Depression increases the risk of mortality among people living with diabetes: Results from national health and nutrition examination survey, USA. Diabetes & Metabolic Syndrome: Clinical Research & Reviews 17, 102892. 10.1016/j.dsx.2023.102892

Kim, D.H., Jensen, A., Jones, K., Raghavan, S., Phillips, L.S., Hung, A., Sun, Y.V., Li, G., Reaven, P., Zhou, H., Zhou, J.J., 2023. A platform for phenotyping disease progression and associated longitudinal risk factors in large-scale EHRs, with application to incident diabetes complications in the UK Biobank. JAMIA Open 6, ooad006. 10.1093/jamiaopen/ooad006

Klarin, D., Lynch, J., Aragam, K., Chaffin, M., Assimes, T.L., Huang, J., Lee, K.M., Shao, Q., Huffman, J.E., Natarajan, P., Arya, S., Small, A., Sun, Y.V., Vujkovic, M., Freiberg, M.S., Wang, L., Chen, J., Saleheen, D., Lee, J.S., Miller, D.R., Reaven, P., Alba, P.R., Patterson, O.V., DuVall, S.L., Boden, W.E., Beckman, J.A., Gaziano, J.M., Concato, J., Rader, D.J., Cho, K., Chang, K.-M., Wilson, P.W.F., O’Donnell, C.J., Kathiresan, S., VA Million Veteran Program, Tsao, P.S., Damrauer, S.M., 2019. Genome-wide association study of peripheral artery disease in the Million Veteran Program. Nat Med 25, 1274–1279. 10.1038/s41591-019-0492-5

Laiteerapong, N., Karter, A.J., Moffet, H.H., Cooper, J.M., Gibbons, R.D., Liu, J.Y., Gao, Y., Huang, E.S., 2017. Ten-year hemoglobin A1c trajectories and outcomes in type 2 diabetes mellitus: The Diabetes & Aging Study. J Diabetes Complications 31, 94–100. 10.1016/j.jdiacomp.2016.07.023

Lennon, H., Kelly, S., Sperrin, M., Buchan, I., Cross, A.J., Leitzmann, M., Cook, M.B., Renehan, A.G., 2018. Framework to construct and interpret latent class trajectory modelling. BMJ Open 8, e020683. 10.1136/bmjopen-2017-020683

Lo, Y., Mendell, N.R., Rubin, D.B., 2001. Testing the number of components in a normal mixture. Biometrika 88, 767–778. 10.1093/biomet/88.3.767

Mésidor, M., Rousseau, M.-C., O’Loughlin, J., Sylvestre, M.-P., 2022. Does group-based trajectory modeling estimate spurious trajectories? BMC Medical Research Methodology 22, 194. 10.1186/s12874-022-01622-9

Nagin, D.S., Jones, B.L., Passos, V.L., Tremblay, R.E., 2018. Group-based multi-trajectory modeling. Stat Methods Med Res 27, 2015–2023. 10.1177/0962280216673085

Nefs, G., Pouwer, F., Denollet, J., Pop, V., 2012. The course of depressive symptoms in primary care patients with type 2 diabetes: results from the Diabetes, Depression, Type D Personality Zuidoost-Brabant (DiaDDZoB) Study. Diabetologia 55, 608–616. 10.1007/s00125-011-2411-2

Noordam, R., Läll, K., Smit, R.A.J., Laisk, T., Metspalu, A., Esko, T., Milani, L., Loos, R.J.F., Mägi, R., Willems van Dijk, K., van Heemst, D., 2021. Stratification of Type 2 Diabetes by Age of Diagnosis in the UK Biobank Reveals Subgroup-Specific Genetic Associations and Causal Risk Profiles. Diabetes 70, 1816–1825. 10.2337/db20-0602

Ong, K.L., Stafford, L.K., McLaughlin, S.A., Boyko, E.J., Vollset, S.E., Smith, A.E., Dalton, B.E., Duprey, J., Cruz, J.A., Hagins, H., Lindstedt, P.A., Aali, A., Abate, Y.H., Abate, M.D., Abbasian, M., Abbasi-Kangevari, Z., Abbasi-Kangevari, M., Abd ElHafeez, S., Abd-Rabu, R., Abdulah, D.M., Abdullah, A.Y.M., Abedi, V., Abidi, H., Aboagye, R.G., Abolhassani, H., Abu-Gharbieh, E., Abu-Zaid, A., Adane, T.D., Adane, D.E., Addo, I.Y., Adegboye, O.A., Adekanmbi, V., Adepoju, A.V., Adnani, Q.E.S., Afolabi, R.F., Agarwal, G., Aghdam, Z.B., Agudelo-Botero, M., Aguilera Arriagada, C.E., Agyemang-Duah, W., Ahinkorah, B.O., Ahmad, D., Ahmad, R., Ahmad, S., Ahmad, A., Ahmadi, A., Ahmadi, K., Ahmed, Ayman, Ahmed, Ali, Ahmed, L.A., Ahmed, S.A., Ajami, M., Akinyemi, R.O., Al Hamad, H., Al Hasan, S.M., AL-Ahdal, T.M.A., Alalwan, T.A., Al-Aly, Z., AlBataineh, M.T., Alcalde-Rabanal, J.E., Alemi, S., Ali, H., Alinia, T., Aljunid, S.M., Almustanyir, S., Al-Raddadi, R.M., Alvis-Guzman, N., Amare, F., Ameyaw, E.K., Amiri, S., Amusa, G.A., Andrei, C.L., Anjana, R.M., Ansar, A., Ansari, G., Ansari-Moghaddam, A., Anyasodor, A.E., Arabloo, J., Aravkin, A.Y., Areda, D., Arifin, H., Arkew, M., Armocida, B., Ärnlöv, J., Artamonov, A.A., Arulappan, J., Aruleba, R.T., Arumugam, A., Aryan, Z., Asemu, M.T., Asghari-Jafarabadi, M., Askari, E., Asmelash, D., Astell-Burt, T., Athar, M., Athari, S.S., Atout, M.M.W., Avila-Burgos, L., Awaisu, A., Azadnajafabad, S., B, D.B., Babamohamadi, H., Badar, M., Badawi, A., Badiye, A.D., Baghcheghi, N., Bagheri, N., Bagherieh, S., Bah, S., Bahadory, S., Bai, R., Baig, A.A., Baltatu, O.C., Baradaran, H.R., Barchitta, M., Bardhan, M., Barengo, N.C., Bärnighausen, T.W., Barone, M.T.U., Barone-Adesi, F., Barrow, A., Bashiri, H., Basiru, A., Basu, Sanjay, Basu, Saurav, Batiha, A.-M.M., Batra, K., Bayih, M.T., Bayileyegn, N.S., Behnoush, A.H., Bekele, A.B., Belete, M.A., Belgaumi, U.I., Belo, L., Bennett, D.A., Bensenor, I.M., Berhe, K., Berhie, A.Y., Bhaskar, S., Bhat, A.N., Bhatti, J.S., Bikbov, B., Bilal, F., Bintoro, B.S., Bitaraf, S., Bitra, V.R., Bjegovic-Mikanovic, V., Bodolica, V., Boloor, A., Brauer, M., Brazo-Sayavera, J., Brenner, H., Butt, Z.A., Calina, D., Campos, L.A., Campos-Nonato, I.R., Cao, Y., Cao, C., Car, J., Carvalho, M., Castañeda-Orjuela, C.A., Catalá-López, F., Cerin, E., Chadwick, J., Chandrasekar, E.K., Chanie, G.S., Charan, J., Chattu, V.K., Chauhan, K., Cheema, H.A., Chekol Abebe, E., Chen, S., Cherbuin, N., Chichagi, F., Chidambaram, S.B., Cho, W.C.S., Choudhari, S.G., Chowdhury, R., Chowdhury, E.K., Chu, D.-T., Chukwu, I.S., Chung, S.-C., Coberly, K., Columbus, A., Contreras, D., Cousin, E., Criqui, M.H., Cruz-Martins, N., Cuschieri, S., Dabo, B., Dadras, O., Dai, X., Damasceno, A.A.M., Dandona, R., Dandona, L., Das, S., Dascalu, A.M., Dash, N.R., Dashti, M., Dávila-Cervantes, C.A., De la Cruz-Góngora, V., Debele, G.R., Delpasand, K., Demisse, F.W., Demissie, G.D., Deng, X., Denova-Gutiérrez, E., Deo, S.V., Dervišević, E., Desai, H.D., Desale, A.T., Dessie, A.M., Desta, F., Dewan, S.M.R., Dey, S., Dhama, K., Dhimal, M., Diao, N., Diaz, D., Dinu, M., Diress, M., Djalalinia, S., Doan, L.P., Dongarwar, D., dos Santos Figueiredo, F.W., Duncan, B.B., Dutta, S., Dziedzic, A.M., Edinur, H.A., Ekholuenetale, M., Ekundayo, T.C., Elgendy, I.Y., Elhadi, M., El-Huneidi, W., Elmeligy, O.A.A., Elmonem, M.A., Endeshaw, D., Esayas, H.L., Eshetu, H.B., Etaee, F., Fadhil, I., Fagbamigbe, A.F., Fahim, A., Falahi, S., Faris, M.E.M., Farrokhpour, H., Farzadfar, F., Fatehizadeh, A., Fazli, G., Feng, X., Ferede, T.Y., Fischer, F., Flood, D., Forouhari, A., Foroumadi, R., Foroutan Koudehi, M., Gaidhane, A.M., Gaihre, S., Gaipov, A., Galali, Y., Ganesan, B., Garcia-Gordillo, M., Gautam, R.K., Gebrehiwot, M., Gebrekidan, K.G., Gebremeskel, T.G., Getacher, L., Ghadirian, F., Ghamari, S.-H., Ghasemi Nour, M., Ghassemi, F., Golechha, M., Goleij, P., Golinelli, D., Gopalani, S.V., Guadie, H.A., Guan, S.-Y., Gudayu, T.W., Guimarães, R.A., Guled, R.A., Gupta, R., Gupta, K., Gupta, V.B., Gupta, V.K., Gyawali, B., Haddadi, R., Hadi, N.R., Haile, T.G., Hajibeygi, R., Haj-Mirzaian, A., Halwani, R., Hamidi, S., Hankey, G.J., Hannan, M.A., Haque, S., Harandi, H., Harlianto, N.I., Hasan, S.M.M., Hasan, S.S., Hasani, H., Hassanipour, S., Hassen, M.B., Haubold, J., Hayat, K., Heidari, G., Heidari, M., Hessami, K., Hiraike, Y., Holla, R., Hossain, S., Hossain, M.S., Hosseini, M.-S., Hosseinzadeh, M., Hosseinzadeh, H., Huang, J., Huda, M.N., Hussain, S., Huynh, H.-H., Hwang, B.-F., Ibitoye, S.E., Ikeda, N., Ilic, I.M., Ilic, M.D., Inbaraj, L.R., Iqbal, A., Islam, S.M.S., Islam, R.M., Ismail, N.E., Iso, H., Isola, G., Itumalla, R., Iwagami, M., Iwu, C.C.D., Iyamu, I.O., Iyasu, A.N., Jacob, L., Jafarzadeh, A., Jahrami, H., Jain, R., Jaja, C., Jamalpoor, Z., Jamshidi, E., Janakiraman, B., Jayanna, K., Jayapal, S.K., Jayaram, S., Jayawardena, R., Jebai, R., Jeong, W., Jin, Y., Jokar, M., Jonas, J.B., Joseph, N., Joseph, A., Joshua, C.E., Joukar, F., Jozwiak, J.J., Kaambwa, B., Kabir, A., Kabthymer, R.H., Kadashetti, V., Kahe, F., Kalhor, R., Kandel, H., Karanth, S.D., Karaye, I.M., Karkhah, S., Katoto, P.D., Kaur, N., Kazemian, S., Kebede, S.A., Khader, Y.S., Khajuria, H., Khalaji, A., Khan, M.A., Khan, M., Khan, A., Khanal, S., Khatatbeh, M.M., Khater, A.M., Khateri, S., khorashadizadeh, F., Khubchandani, J., Kibret, B.G., Kim, M.S., Kimokoti, R.W., Kisa, A., Kivimäki, M., Kolahi, A.-A., Komaki, S., Kompani, F., Koohestani, H.R., Korzh, O., Kostev, K., Kothari, N., Koyanagi, A., Krishan, K., Krishnamoorthy, Y., Kuate Defo, B., Kuddus, M., Kuddus, M.A., Kumar, R., Kumar, H., Kundu, S., Kurniasari, M.D., Kuttikkattu, A., La Vecchia, C., Lallukka, T., Larijani, B., Larsson, A.O., Latief, K., Lawal, B.K., Le, T.T.T., Le, T.T.B., Lee, S.W.H., Lee, M., Lee, W.-C., Lee, P.H., Lee, S., Lee, S.W., Legesse, S.M., Lenzi, J., Li, Y., Li, M.-C., Lim, S.S., Lim, L.-L., Liu, X., Liu, C., Lo, C.-H., Lopes, G., Lorkowski, S., Lozano, R., Lucchetti, G., Maghazachi, A.A., Mahasha, P.W., Mahjoub, S., Mahmoud, M.A., Mahmoudi, R., Mahmoudimanesh, M., Mai, A.T., Majeed, A., Majma Sanaye, P., Makris, K.C., Malhotra, K., Malik, A.A., Malik, I., Mallhi, T.H., Malta, D.C., Mamun, A.A., Mansouri, B., Marateb, H.R., Mardi, P., Martini, S., Martorell, M., Marzo, R.R., Masoudi, R., Masoudi, S., Mathews, E., Maugeri, A., Mazzaglia, G., Mekonnen, T., Meshkat, M., Mestrovic, T., Miao Jonasson, J., Miazgowski, T., Michalek, I.M., Minh, L.H.N., Mini, G., Miranda, J.J., Mirfakhraie, R., Mirrakhimov, E.M., Mirza-Aghazadeh-Attari, M., Misganaw, A., Misgina, K.H., Mishra, M., Moazen, B., Mohamed, N.S., Mohammadi, E., Mohammadi, M., Mohammadian-Hafshejani, A., Mohammadshahi, M., Mohseni, A., Mojiri-forushani, H., Mokdad, A.H., Momtazmanesh, S., Monasta, L., Moniruzzaman, M., Mons, U., Montazeri, F., Moodi Ghalibaf, A., Moradi, Y., Moradi, M., Moradi Sarabi, M., Morovatdar, N., Morrison, S.D., Morze, J., Mossialos, E., Mostafavi, E., Mueller, U.O., Mulita, F., Mulita, A., Murillo-Zamora, E., Musa, K.I., Mwita, J.C., Nagaraju, S.P., Naghavi, M., Nainu, F., Nair, T.S., Najmuldeen, H.H.R., Nangia, V., Nargus, S., Naser, A.Y., Nassereldine, H., Natto, Z.S., Nauman, J., Nayak, B.P., Ndejjo, R., Negash, H., Negoi, R.I., Nguyen, H.T.H., Nguyen, D.H., Nguyen, P.T., Nguyen, V.T., Nguyen, H.Q., Niazi, R.K., Nigatu, Y.T., Ningrum, D.N.A., Nizam, M.A., Nnyanzi, L.A., Noreen, M., Noubiap, J.J., Nzoputam, O.J., Nzoputam, C.I., Oancea, B., Odogwu, N.M., Odukoya, O.O., Ojha, V.A., Okati-Aliabad, H., Okekunle, A.P., Okonji, O.C., Okwute, P.G., Olufadewa, I.I., Onwujekwe, O.E., Ordak, M., Ortiz, A., Osuagwu, U.L., Oulhaj, A., Owolabi, M.O., Padron-Monedero, A., Padubidri, J.R., Palladino, R., Panagiotakos, D., Panda-Jonas, S., Pandey, Ashok, Pandey, Anamika, Pandi-Perumal, S.R., Pantea Stoian, A.M., Pardhan, S., Parekh, T., Parekh, U., Pasovic, M., Patel, J., Patel, J.R., Paudel, U., Pepito, V.C.F., Pereira, M., Perico, N., Perna, S., Petcu, I.-R., Petermann-Rocha, F.E., Podder, V., Postma, M.J., Pourali, G., Pourtaheri, N., Prates, E.J.S., Qadir, M.M.F., Qattea, I., Raee, P., Rafique, I., Rahimi, M., Rahimifard, M., Rahimi-Movaghar, V., Rahman, M.O., Rahman, M.A., Rahman, M.H.U., Rahman, M., Rahman, M.M., Rahmani, M., Rahmani, S., Rahmanian, V., Rahmawaty, S., Rahnavard, N., Rajbhandari, B., Ram, P., Ramazanu, S., Rana, J., Rancic, N., Ranjha, M.M.A.N., Rao, C.R., Rapaka, D., Rasali, D.P., Rashedi, S., Rashedi, V., Rashid, A.M., Rashidi, M.-M., Ratan, Z.A., Rawaf, S., Rawal, L., Redwan, E.M.M., Remuzzi, G., Rengasamy, K.R., Renzaho, A.M.N., Reyes, L.F., Rezaei, Nima, Rezaei, Nazila, Rezaeian, M., Rezazadeh, H., Riahi, S.M., Rias, Y.A., Riaz, M., Ribeiro, D., Rodrigues, M., Rodriguez, J.A.B., Roever, L., Rohloff, P., Roshandel, G., Roustazadeh, A., Rwegerera, G.M., Saad, A.M.A., Saber-Ayad, M.M., Sabour, S., Sabzmakan, L., Saddik, B., Sadeghi, E., Saeed, U., Saeedi Moghaddam, S., Safi, S., Safi, S.Z., Saghazadeh, A., Saheb Sharif-Askari, N., Saheb Sharif-Askari, F., Sahebkar, A., Sahoo, S.S., Sahoo, H., Saif-Ur-Rahman, K., Sajid, M.R., Salahi, Sarvenaz, Salahi, Saina, Saleh, M.A., Salehi, M.A., Salomon, J.A., Sanabria, J., Sanjeev, R.K., Sanmarchi, F., Santric-Milicevic, M.M., Sarasmita, M.A., Sargazi, S., Sathian, B., Sathish, T., Sawhney, M., Schlaich, M.P., Schmidt, M.I., Schuermans, A., Seidu, A.-A., Senthil Kumar, N., Sepanlou, S.G., Sethi, Y., Seylani, A., Shabany, M., Shafaghat, T., Shafeghat, M., Shafie, M., Shah, N.S., Shahid, S., Shaikh, M.A., Shanawaz, M., Shannawaz, M., Sharfaei, S., Shashamo, B.B., Shiri, R., Shittu, A., Shivakumar, K.M., Shivalli, S., Shobeiri, P., Shokri, F., Shuval, K., Sibhat, M.M., Silva, L.M.L.R., Simpson, C.R., Singh, J.A., Singh, P., Singh, S., Siraj, M.S., Skryabina, A.A., Sohag, A.A.M., Soleimani, H., Solikhah, S., Soltani-Zangbar, M.S., Somayaji, R., Sorensen, R.J.D., Starodubova, A.V., Sujata, S., Suleman, M., Sun, J., Sundström, J., Tabarés-Seisdedos, R., Tabatabaei, S.M., Tabatabaeizadeh, S.-A., Tabish, M., Taheri, M., Taheri, E., Taki, E., Tamuzi, J.J.L., Tan, K.-K., Tat, N.Y., Taye, B.T., Temesgen, W.A., Temsah, M.-H., Tesler, R., Thangaraju, P., Thankappan, K.R., Thapa, R., Tharwat, S., Thomas, N., Ticoalu, J.H.V., Tiyuri, A., Tonelli, M., Tovani-Palone, M.R., Trico, D., Trihandini, I., Tripathy, J.P., Tromans, S.J., Tsegay, G.M., Tualeka, A.R., Tufa, D.G., Tyrovolas, S., Ullah, S., Upadhyay, E., Vahabi, S.M., Vaithinathan, A.G., Valizadeh, R., van Daalen, K.R., Vart, P., Varthya, S.B., Vasankari, T.J., Vaziri, S., Verma, M. verma, Verras, G.-I., Vo, D.C., Wagaye, B., Waheed, Y., Wang, Z., Wang, Y., Wang, C., Wang, F., Wassie, G.T., Wei, M.Y.W., Weldemariam, A.H., Westerman, R., Wickramasinghe, N.D., Wu, Y., Wulandari, R.D., Xia, J., Xiao, H., Xu, S., Xu, X., Yada, D.Y., Yang, L., Yatsuya, H., Yesiltepe, M., Yi, S., Yohannis, H.K., Yonemoto, N., You, Y., Zaman, S.B., Zamora, N., Zare, I., Zarea, K., Zarrintan, A., Zastrozhin, M.S., Zeru, N.G., Zhang, Z.-J., Zhong, C., Zhou, J., Zielińska, M., Zikarg, Y.T., Zodpey, S., Zoladl, M., Zou, Z., Zumla, A., Zuniga, Y.M.H., Magliano, D.J., Murray, C.J.L., Hay, S.I., Vos, T., 2023. Global, regional, and national burden of diabetes from 1990 to 2021, with projections of prevalence to 2050: a systematic analysis for the Global Burden of Disease Study 2021. The Lancet 402, 203–234. 10.1016/S0140-6736(23)01301-6

Pain, O., Al-Chalabi, A., Lewis, C.M., 2024. The GenoPred Pipeline: A Comprehensive and Scalable Pipeline for Polygenic Scoring. 10.1101/2024.06.12.24308843

Pei, J., Wang, X., Pei, Z., Hu, X., 2023. Glycemic control, HbA1c variability, and major cardiovascular adverse outcomes in type 2 diabetes patients with elevated cardiovascular risk: insights from the ACCORD study. Cardiovasc Diabetol 22, 287. 10.1186/s12933-023-02026-9

Proust-Lima, C., Philipps, V., Liquet, B., 2017. Estimation of Extended Mixed Models Using Latent Classes and Latent Processes: The R Package lcmm. Journal of Statistical Software 78, 1–56. 10.18637/jss.v078.i02

Ram, N., Grimm, K.J., 2009. Growth Mixture Modeling: A Method for Identifying Differences in Longitudinal Change Among Unobserved Groups. Int J Behav Dev 33, 565–576. 10.1177/0165025409343765

Ripley, B., 2009. nnet: Feed-Forward Neural Networks and Multinomial Log-Linear Models. 10.32614/CRAN.package.nnet

Rodriguez, P., San Martin, V.T., Pantalone, K.M., 2024. Therapeutic Inertia in the Management of Type 2 Diabetes: A Narrative Review. Diabetes Ther 15, 567–583. 10.1007/s13300-024-01530-9

Shahabi, N., Fakhri, Y., Aghamolaei, T., Hosseini, Z., Homayuni, A., 2023. Socio-personal factors affecting adherence to treatment in patients with type 2 diabetes: A systematic review and meta-analysis. Primary Care Diabetes 17, 205–220. 10.1016/j.pcd.2023.03.005

Spanakis, E.K., Golden, S.H., 2013. Race/ethnic difference in diabetes and diabetic complications. Curr Diab Rep 13, 814–823. 10.1007/s11892-013-0421-9

Sudlow, C., Gallacher, J., Allen, N., Beral, V., Burton, P., Danesh, J., Downey, P., Elliott, P., Green, J., Landray, M., Liu, B., Matthews, P., Ong, G., Pell, J., Silman, A., Young, A., Sprosen, T., Peakman, T., Collins, R., 2015. UK Biobank: An Open Access Resource for Identifying the Causes of a Wide Range of Complex Diseases of Middle and Old Age. PLOS Medicine 12, e1001779. 10.1371/journal.pmed.1001779

Therneau, T., Gambsch, P., n.d. Modeling Survival Data: Extending the Cox Model | SpringerLink [WWW Document]. URL https://link.springer.com/book/10.1007/978-1-4757-3294-8 (accessed 9.18.24).

Vasishta, S., Ganesh, K., Umakanth, S., Joshi, M.B., 2022. Ethnic disparities attributed to the manifestation in and response to type 2 diabetes: insights from metabolomics. Metabolomics 18, 45. 10.1007/s11306-022-01905-8

Vithian, K., Hurel, S., 2010. Microvascular complications: pathophysiology and management. Clin Med (Lond) 10, 505–509. 10.7861/clinmedicine.10-5-505

Wickham, H., 2016. ggplot2, Use R! Springer International Publishing, Cham. 10.1007/978-3-319-24277-4

Wilmot, E., Idris, I., 2014. Early onset type 2 diabetes: risk factors, clinical impact and management. Therapeutic Advances in Chronic Disease 5, 234. 10.1177/2040622314548679

Xu, F., Earp, J.E., Riebe, D., Delmonico, M.J., Lofgren, I.E., Greene, G.W., 2024. The relationship between fat distribution and diabetes in US adults by race/ethnicity. Front. Public Health 12. 10.3389/fpubh.2024.1373544

Young, K.G., McDonald, T.J., Shields, B.M., 2022. Glycated haemoglobin measurements from UK Biobank are different to those in linked primary care records: implications for combining biochemistry data from research studies and routine clinical care. International Journal of Epidemiology 51, 1022–1024. 10.1093/ije/dyab265

Yu, M., Zhang, X., Lu, F., Fang, L., 2015. Depression and Risk for Diabetes: A Meta-Analysis. Can J Diabetes 39, 266–272. 10.1016/j.jcjd.2014.11.006

