## Supplementary Information for "Latent class growth mixture modelling of HbA1C trajectories identifies individuals at high risk of developing complications of type 2 diabetes mellitus in the UK Biobank"

**Methods S1. Sample quality control**

Standard sample quality control for the entire UKB cohort was applied using previously described methods (Bycroft *et al*., 2018). Relatedness was determined using the GreedyRelated R package. Genetic ancestry was determined using the UKBKings R package, which uses a previously validated method for ancestral determination (Pain e*t al*., 2024).

**Methods S2. LCGMM specification and computation time**

The Latent Class Growth Mixture Model was generated using the default variance-covariance structure applied by the *lcmm* R package (Proust-Lima *et al*., 2017). The model was allowed a maximum of 200 iterations for convergence, and 100 random start points for the model were used to avoid solutions to avoid local maxima (Hipp & Bauer, 2006). Models were computed in parallel using 50 CPU cores, and a maximum of 48 hours was allowed for convergence. The six-class solution took 42.7 hours to converge, and the seven-class solution did not converge within the allotted time frame.

**Methods S3. Polygenic scores generation**

PRSCS, via GenoPredPipe was used to Polygenic scores for three traits: Body mass index, Type 2 diabetes (T2D), and major depressive disorder (MDD) using GWAS summary statistics containing individuals of European ancestry that had little sample overlap with the UK Biobank (Locke *et al*., 2015; Scott *et al*., 2017; Wray *et al*., 2018; Ge *et* al., 2019; Pain *et al*., 2024).

**Methods S4. Polygenic score validation**

Polygenic score validation was performed in all available unrelated individuals in the UK Biobank of European ancestry for BMI, and in individuals who met these criteria and had primary care record data available for T2D and MDD.

BMI was defined using UK Biobank field 21001. T2D status was derived using the cohort outlined by the primary text, and Individuals who did not meet any criteria for T2D diagnosis were controls. MDD was derived solely in individuals with primary care record data available, according to previously described methods (Fabbri *et al*., 2021).

Linear regression was used for BMI, and logistic regression was used for T2D status and MDD status, to determine the association of the polygenic scores with their respective traits.

**Results S1. Polygenic score validation**

All three polygenic scores were robustly associated with their respective outcome traits (Supplementary information, Table 1).

Supplementary Information, Table 1. The association of polygenic scores with their relevant traits. R^2^ is reported for linear regression, and Nagelkerke’s pseudo-R^2^ is reported for logistic regression.

| **Polygenic score** | **R^2^** | **p-value** |
| --- | --- | --- |
| Body mass index | 0.081 | < 1x10^-200^ |
| Type 2 diabetes mellitus | 0.040 | < 1x10^-200^ |
| Major depressive disorder | 0.0073 | 5.59x10^-156^ |

**References**

Bycroft, C., Freeman, C., Petkova, D., Band, G., Elliott, L.T., Sharp, K., Motyer, A., Vukcevic, D., Delaneau, O., O’Connell, J., Cortes, A., Welsh, S., Young, A., Effingham, M., McVean, G., Leslie, S., Allen, N., Donnelly, P., Marchini, J., 2018. The UK Biobank resource with deep phenotyping and genomic data. Nature 562, 203–209. <https://doi.org/10.1038/s41586-018-0579-z>

Fabbri, C., Hagenaars, S.P., John, C., Williams, A.T., Shrine, N., Moles, L., Hanscombe, K.B., Serretti, A., Shepherd, D.J., Free, R.C., Wain, L.V., Tobin, M.D., Lewis, C.M., 2021. Genetic and clinical characteristics of treatment-resistant depression using primary care records in two UK cohorts. Mol Psychiatry 26, 3363–3373. <https://doi.org/10.1038/s41380-021-01062-9>

Ge, T., Chen, C.-Y., Ni, Y., Feng, Y.-C.A., Smoller, J.W., 2019. Polygenic prediction via Bayesian regression and continuous shrinkage priors. Nat Commun 10, 1776. <https://doi.org/10.1038/s41467-019-09718-5>

Hipp, J.R., Bauer, D.J., 2006. “Local Solutions in the Estimation of Growth Mixture Models”: Correction to Hipp and Bauer (2006). Psychological Methods 11, 305–305. <https://doi.org/10.1037/1082-989X.11.3.305>

Locke, A.E., Kahali, B., Berndt, S.I., Justice, A.E., Pers, T.H., Day, F.R., Powell, C., Vedantam, S., Buchkovich, M.L., Yang, J., Croteau-Chonka, D.C., Esko, T., Fall, T., Ferreira, T., Gustafsson, S., Kutalik, Z., Luan, J., Mägi, R., Randall, J.C., Winkler, T.W., Wood, A.R., Workalemahu, T., Faul, J.D., Smith, J.A., Hua Zhao, J., Zhao, W., Chen, J., Fehrmann, R., Hedman, Å.K., Karjalainen, J., Schmidt, E.M., Absher, D., Amin, N., Anderson, D., Beekman, M., Bolton, J.L., Bragg-Gresham, J.L., Buyske, S., Demirkan, A., Deng, G., Ehret, G.B., Feenstra, B., Feitosa, M.F., Fischer, K., Goel, A., Gong, J., Jackson, A.U., Kanoni, S., Kleber, M.E., Kristiansson, K., Lim, U., Lotay, V., Mangino, M., Mateo Leach, I., Medina-Gomez, C., Medland, S.E., Nalls, M.A., Palmer, C.D., Pasko, D., Pechlivanis, S., Peters, M.J., Prokopenko, I., Shungin, D., Stančáková, A., Strawbridge, R.J., Ju Sung, Y., Tanaka, Toshiko, Teumer, A., Trompet, S., van der Laan, S.W., van Setten, J., Van Vliet-Ostaptchouk, J.V., Wang, Z., Yengo, L., Zhang, W., Isaacs, A., Albrecht, E., Ärnlöv, J., Arscott, G.M., Attwood, A.P., Bandinelli, S., Barrett, A., Bas, I.N., Bellis, C., Bennett, A.J., Berne, C., Blagieva, R., Blüher, M., Böhringer, S., Bonnycastle, L.L., Böttcher, Y., Boyd, H.A., Bruinenberg, M., Caspersen, I.H., Ida Chen, Y.-D., Clarke, R., Warwick Daw, E., de Craen, A.J.M., Delgado, G., Dimitriou, M., Doney, A.S.F., Eklund, N., Estrada, K., Eury, E., Folkersen, L., Fraser, R.M., Garcia, M.E., Geller, F., Giedraitis, V., Gigante, B., Go, A.S., Golay, A., Goodall, A.H., Gordon, S.D., Gorski, M., Grabe, H.-J., Grallert, H., Grammer, T.B., Gräßler, J., Grönberg, H., Groves, C.J., Gusto, G., Haessler, J., Hall, P., Haller, T., Hallmans, G., Hartman, C.A., Hassinen, M., Hayward, C., Heard-Costa, N.L., Helmer, Q., Hengstenberg, C., Holmen, O., Hottenga, J.-J., James, A.L., Jeff, J.M., Johansson, Å., Jolley, J., Juliusdottir, T., Kinnunen, L., Koenig, W., Koskenvuo, M., Kratzer, W., Laitinen, J., Lamina, C., Leander, K., Lee, N.R., Lichtner, P., Lind, L., Lindström, J., Sin Lo, K., Lobbens, S., Lorbeer, R., Lu, Y., Mach, F., Magnusson, P.K.E., Mahajan, A., McArdle, W.L., McLachlan, S., Menni, C., Merger, S., Mihailov, E., Milani, L., Moayyeri, A., Monda, K.L., Morken, M.A., Mulas, A., Müller, G., Müller-Nurasyid, M., Musk, A.W., Nagaraja, R., Nöthen, M.M., Nolte, I.M., Pilz, S., Rayner, N.W., Renstrom, F., Rettig, R., Ried, J.S., Ripke, S., Robertson, N.R., Rose, L.M., Sanna, S., Scharnagl, H., Scholtens, S., Schumacher, F.R., Scott, W.R., Seufferlein, T., Shi, J., Vernon Smith, A., Smolonska, J., Stanton, A.V., Steinthorsdottir, V., Stirrups, K., Stringham, H.M., Sundström, J., Swertz, M.A., Swift, A.J., Syvänen, A.-C., Tan, S.-T., Tayo, B.O., Thorand, B., Thorleifsson, G., Tyrer, J.P., Uh, H.-W., Vandenput, L., Verhulst, F.C., Vermeulen, S.H., Verweij, N., Vonk, J.M., Waite, L.L., Warren, H.R., Waterworth, D., Weedon, M.N., Wilkens, L.R., Willenborg, C., Wilsgaard, T., Wojczynski, M.K., Wong, A., Wright, A.F., Zhang, Q., Brennan, E.P., Choi, M., Dastani, Z., Drong, A.W., Eriksson, P., Franco-Cereceda, A., Gådin, J.R., Gharavi, A.G., Goddard, M.E., Handsaker, R.E., Huang, J., Karpe, F., Kathiresan, S., Keildson, S., Kiryluk, K., Kubo, M., Lee, J.-Y., Liang, L., Lifton, R.P., Ma, B., McCarroll, S.A., McKnight, A.J., Min, J.L., Moffatt, M.F., Montgomery, G.W., Murabito, J.M., Nicholson, G., Nyholt, D.R., Okada, Y., Perry, J.R.B., Dorajoo, R., Reinmaa, E., Salem, R.M., Sandholm, N., Scott, R.A., Stolk, L., Takahashi, A., Tanaka, Toshihiro, van’t Hooft, F.M., Vinkhuyzen, A.A.E., Westra, H.-J., Zheng, W., Zondervan, K.T., Heath, A.C., Arveiler, D., Bakker, S.J.L., Beilby, J., Bergman, R.N., Blangero, J., Bovet, P., Campbell, H., Caulfield, M.J., Cesana, G., Chakravarti, A., Chasman, D.I., Chines, P.S., Collins, F.S., Crawford, D.C., Adrienne Cupples, L., Cusi, D., Danesh, J., de Faire, U., den Ruijter, H.M., Dominiczak, A.F., Erbel, R., Erdmann, J., Eriksson, J.G., Farrall, M., Felix, S.B., Ferrannini, E., Ferrières, J., Ford, I., Forouhi, N.G., Forrester, T., Franco, O.H., Gansevoort, R.T., Gejman, P.V., Gieger, C., Gottesman, O., Gudnason, V., Gyllensten, U., Hall, A.S., Harris, T.B., Hattersley, A.T., Hicks, A.A., Hindorff, L.A., Hingorani, A.D., Hofman, A., Homuth, G., Kees Hovingh, G., Humphries, S.E., Hunt, S.C., Hyppönen, E., Illig, T., Jacobs, K.B., Jarvelin, M.-R., Jöckel, K.-H., Johansen, B., Jousilahti, P., Wouter Jukema, J., Jula, A.M., Kaprio, J., Kastelein, J.J.P., Keinanen-Kiukaanniemi, S.M., Kiemeney, L.A., Knekt, P., Kooner, J.S., Kooperberg, C., Kovacs, P., Kraja, A.T., Kumari, M., Kuusisto, J., Lakka, T.A., Langenberg, C., Le Marchand, L., Lehtimäki, T., Lyssenko, V., Männistö, S., Marette, A., Matise, T.C., McKenzie, C.A., McKnight, B., Moll, F.L., Morris, A.D., Morris, A.P., Murray, J.C., Nelis, M., Ohlsson, C., Oldehinkel, A.J., Ong, K.K., Madden, P.A.F., Pasterkamp, G., Peden, J.F., Peters, A., Postma, D.S., Pramstaller, P.P., Price, J.F., Qi, L., Raitakari, O.T., Rankinen, T., Rao, D.C., Rice, T.K., Ridker, P.M., Rioux, J.D., Ritchie, M.D., Rudan, I., Salomaa, V., Samani, N.J., Saramies, J., Sarzynski, M.A., Schunkert, H., Schwarz, P.E.H., Sever, P., Shuldiner, A.R., Sinisalo, J., Stolk, R.P., Strauch, K., Tönjes, A., Trégouët, D.-A., Tremblay, A., Tremoli, E., Virtamo, J., Vohl, M.-C., Völker, U., Waeber, G., Willemsen, G., Witteman, J.C., Carola Zillikens, M., Adair, L.S., Amouyel, P., Asselbergs, F.W., Assimes, T.L., Bochud, M., Boehm, B.O., Boerwinkle, E., Bornstein, S.R., Bottinger, E.P., Bouchard, C., Cauchi, S., Chambers, J.C., Chanock, S.J., Cooper, R.S., de Bakker, P.I.W., Dedoussis, G., Ferrucci, L., Franks, P.W., Froguel, P., Groop, L.C., Haiman, C.A., Hamsten, A., Hui, J., Hunter, D.J., Hveem, K., Kaplan, R.C., Kivimaki, M., Kuh, D., Laakso, M., Liu, Y., Martin, N.G., März, W., Melbye, M., Metspalu, A., Moebus, S., Munroe, P.B., Njølstad, I., Oostra, B.A., Palmer, C.N.A., Pedersen, N.L., Perola, M., Pérusse, L., Peters, U., Power, C., Quertermous, T., Rauramaa, R., Rivadeneira, F., Saaristo, T.E., Saleheen, D., Sattar, N., Schadt, E.E., Schlessinger, D., Eline Slagboom, P., Snieder, H., Spector, T.D., Thorsteinsdottir, U., Stumvoll, M., Tuomilehto, J., Uitterlinden, A.G., Uusitupa, M., van der Harst, P., Walker, M., Wallaschofski, H., Wareham, N.J., Watkins, H., Weir, D.R., Wichmann, H.-E., Wilson, J.F., Zanen, P., Borecki, I.B., Deloukas, P., Fox, C.S., Heid, I.M., O’Connell, J.R., Strachan, D.P., Stefansson, K., van Duijn, C.M., Abecasis, G.R., Franke, L., Frayling, T.M., McCarthy, M.I., Visscher, P.M., Scherag, A., Willer, C.J., Boehnke, M., Mohlke, K.L., Lindgren, C.M., Beckmann, J.S., Barroso, I., North, K.E., Ingelsson, E., Hirschhorn, J.N., Loos, R.J.F., Speliotes, E.K., 2015. Genetic studies of body mass index yield new insights for obesity biology. Nature 518, 197–206. <https://doi.org/10.1038/nature14177>

Pain, O., Al-Chalabi, A., Lewis, C.M., 2024. The GenoPred Pipeline: A Comprehensive and Scalable Pipeline for Polygenic Scoring. <https://doi.org/10.1101/2024.06.12.24308843>

Proust-Lima, C., Philipps, V., Liquet, B., 2017. Estimation of Extended Mixed Models Using Latent Classes and Latent Processes: The R Package lcmm. Journal of Statistical Software 78, 1–56. <https://doi.org/10.18637/jss.v078.i02>

Scott, R.A., Scott, L.J., Mägi, R., Marullo, L., Gaulton, K.J., Kaakinen, M., Pervjakova, N., Pers, T.H., Johnson, A.D., Eicher, J.D., Jackson, A.U., Ferreira, T., Lee, Y., Ma, C., Steinthorsdottir, V., Thorleifsson, G., Qi, L., Van Zuydam, N.R., Mahajan, A., Chen, H., Almgren, P., Voight, B.F., Grallert, H., Müller-Nurasyid, M., Ried, J.S., Rayner, N.W., Robertson, N., Karssen, L.C., van Leeuwen, E.M., Willems, S.M., Fuchsberger, C., Kwan, P., Teslovich, T.M., Chanda, P., Li, M., Lu, Y., Dina, C., Thuillier, D., Yengo, L., Jiang, L., Sparso, T., Kestler, H.A., Chheda, H., Eisele, L., Gustafsson, S., Frånberg, M., Strawbridge, R.J., Benediktsson, R., Hreidarsson, A.B., Kong, A., Sigurðsson, G., Kerrison, N.D., Luan, J., Liang, L., Meitinger, T., Roden, M., Thorand, B., Esko, T., Mihailov, E., Fox, C., Liu, C.-T., Rybin, D., Isomaa, B., Lyssenko, V., Tuomi, T., Couper, D.J., Pankow, J.S., Grarup, N., Have, C.T., Jørgensen, M.E., Jørgensen, T., Linneberg, A., Cornelis, M.C., van Dam, R.M., Hunter, D.J., Kraft, P., Sun, Q., Edkins, S., Owen, K.R., Perry, J.R.B., Wood, A.R., Zeggini, E., Tajes-Fernandes, J., Abecasis, G.R., Bonnycastle, L.L., Chines, P.S., Stringham, H.M., Koistinen, H.A., Kinnunen, L., Sennblad, B., Mühleisen, T.W., Nöthen, M.M., Pechlivanis, S., Baldassarre, D., Gertow, K., Humphries, S.E., Tremoli, E., Klopp, N., Meyer, J., Steinbach, G., Wennauer, R., Eriksson, J.G., Mӓnnistö, S., Peltonen, L., Tikkanen, E., Charpentier, G., Eury, E., Lobbens, S., Gigante, B., Leander, K., McLeod, O., Bottinger, E.P., Gottesman, O., Ruderfer, D., Blüher, M., Kovacs, P., Tonjes, A., Maruthur, N.M., Scapoli, C., Erbel, R., Jöckel, K.-H., Moebus, S., de Faire, U., Hamsten, A., Stumvoll, M., Deloukas, P., Donnelly, P.J., Frayling, T.M., Hattersley, A.T., Ripatti, S., Salomaa, V., Pedersen, N.L., Boehm, B.O., Bergman, R.N., Collins, F.S., Mohlke, K.L., Tuomilehto, J., Hansen, T., Pedersen, O., Barroso, I., Lannfelt, L., Ingelsson, E., Lind, L., Lindgren, C.M., Cauchi, S., Froguel, P., Loos, R.J.F., Balkau, B., Boeing, H., Franks, P.W., Barricarte Gurrea, A., Palli, D., van der Schouw, Y.T., Altshuler, D., Groop, L.C., Langenberg, C., Wareham, N.J., Sijbrands, E., van Duijn, C.M., Florez, J.C., Meigs, J.B., Boerwinkle, E., Gieger, C., Strauch, K., Metspalu, A., Morris, A.D., Palmer, C.N.A., Hu, F.B., Thorsteinsdottir, U., Stefansson, K., Dupuis, J., Morris, A.P., Boehnke, M., McCarthy, M.I., Prokopenko, I., DIAbetes Genetics Replication And Meta-analysis (DIAGRAM) Consortium, 2017. An Expanded Genome-Wide Association Study of Type 2 Diabetes in Europeans. Diabetes 66, 2888–2902. <https://doi.org/10.2337/db16-1253>

Wray, N.R., Ripke, S., Mattheisen, M., Trzaskowski, M., Byrne, E.M., Abdellaoui, A., Adams, M.J., Agerbo, E., Air, T.M., Andlauer, T.M.F., Bacanu, S.-A., Bækvad-Hansen, M., Beekman, A.F.T., Bigdeli, T.B., Binder, E.B., Blackwood, D.R.H., Bryois, J., Buttenschøn, H.N., Bybjerg-Grauholm, J., Cai, N., Castelao, E., Christensen, J.H., Clarke, T.-K., Coleman, J.I.R., Colodro-Conde, L., Couvy-Duchesne, B., Craddock, N., Crawford, G.E., Crowley, C.A., Dashti, H.S., Davies, G., Deary, I.J., Degenhardt, F., Derks, E.M., Direk, N., Dolan, C.V., Dunn, E.C., Eley, T.C., Eriksson, N., Escott-Price, V., Kiadeh, F.H.F., Finucane, H.K., Forstner, A.J., Frank, J., Gaspar, H.A., Gill, M., Giusti-Rodríguez, P., Goes, F.S., Gordon, S.D., Grove, J., Hall, L.S., Hannon, E., Hansen, C.S., Hansen, T.F., Herms, S., Hickie, I.B., Hoffmann, P., Homuth, G., Horn, C., Hottenga, J.-J., Hougaard, D.M., Hu, M., Hyde, C.L., Ising, M., Jansen, R., Jin, F., Jorgenson, E., Knowles, J.A., Kohane, I.S., Kraft, J., Kretzschmar, W.W., Krogh, J., Kutalik, Z., Lane, J.M., Li, Yihan, Li, Yun, Lind, P.A., Liu, X., Lu, L., MacIntyre, D.J., MacKinnon, D.F., Maier, R.M., Maier, W., Marchini, J., Mbarek, H., McGrath, P., McGuffin, P., Medland, S.E., Mehta, D., Middeldorp, C.M., Mihailov, E., Milaneschi, Y., Milani, L., Mill, J., Mondimore, F.M., Montgomery, G.W., Mostafavi, S., Mullins, N., Nauck, M., Ng, B., Nivard, M.G., Nyholt, D.R., O’Reilly, P.F., Oskarsson, H., Owen, M.J., Painter, J.N., Pedersen, C.B., Pedersen, M.G., Peterson, R.E., Pettersson, E., Peyrot, W.J., Pistis, G., Posthuma, D., Purcell, S.M., Quiroz, J.A., Qvist, P., Rice, J.P., Riley, B.P., Rivera, M., Saeed Mirza, S., Saxena, R., Schoevers, R., Schulte, E.C., Shen, L., Shi, J., Shyn, S.I., Sigurdsson, E., Sinnamon, G.B.C., Smit, J.H., Smith, D.J., Stefansson, H., Steinberg, S., Stockmeier, C.A., Streit, F., Strohmaier, J., Tansey, K.E., Teismann, H., Teumer, A., Thompson, W., Thomson, P.A., Thorgeirsson, T.E., Tian, C., Traylor, M., Treutlein, J., Trubetskoy, V., Uitterlinden, A.G., Umbricht, D., Van der Auwera, S., van Hemert, A.M., Viktorin, A., Visscher, P.M., Wang, Y., Webb, B.T., Weinsheimer, S.M., Wellmann, J., Willemsen, G., Witt, S.H., Wu, Y., Xi, H.S., Yang, J., Zhang, F., Arolt, V., Baune, B.T., Berger, K., Boomsma, D.I., Cichon, S., Dannlowski, U., de Geus, E.C.J., DePaulo, J.R., Domenici, E., Domschke, K., Esko, T., Grabe, H.J., Hamilton, S.P., Hayward, C., Heath, A.C., Hinds, D.A., Kendler, K.S., Kloiber, S., Lewis, G., Li, Q.S., Lucae, S., Madden, P.F.A., Magnusson, P.K., Martin, N.G., McIntosh, A.M., Metspalu, A., Mors, O., Mortensen, P.B., Müller-Myhsok, B., Nordentoft, M., Nöthen, M.M., O’Donovan, M.C., Paciga, S.A., Pedersen, N.L., Penninx, B.W.J.H., Perlis, R.H., Porteous, D.J., Potash, J.B., Preisig, M., Rietschel, M., Schaefer, C., Schulze, T.G., Smoller, J.W., Stefansson, K., Tiemeier, H., Uher, R., Völzke, H., Weissman, M.M., Werge, T., Winslow, A.R., Lewis, C.M., Levinson, D.F., Breen, G., Børglum, A.D., Sullivan, P.F., 2018. Genome-wide association analyses identify 44 risk variants and refine the genetic architecture of major depression. Nat Genet 50, 668–681. <https://doi.org/10.1038/s41588-018-0090-3>
