## Supplementary figures and images for "Latent class growth mixture modelling of HbA1C trajectories identifies individuals at high risk of developing complications of type 2 diabetes mellitus in the UK Biobank"

### Supplementary Figure 1

Figure 1. Upset plot of T2D diagnostic criteria met.

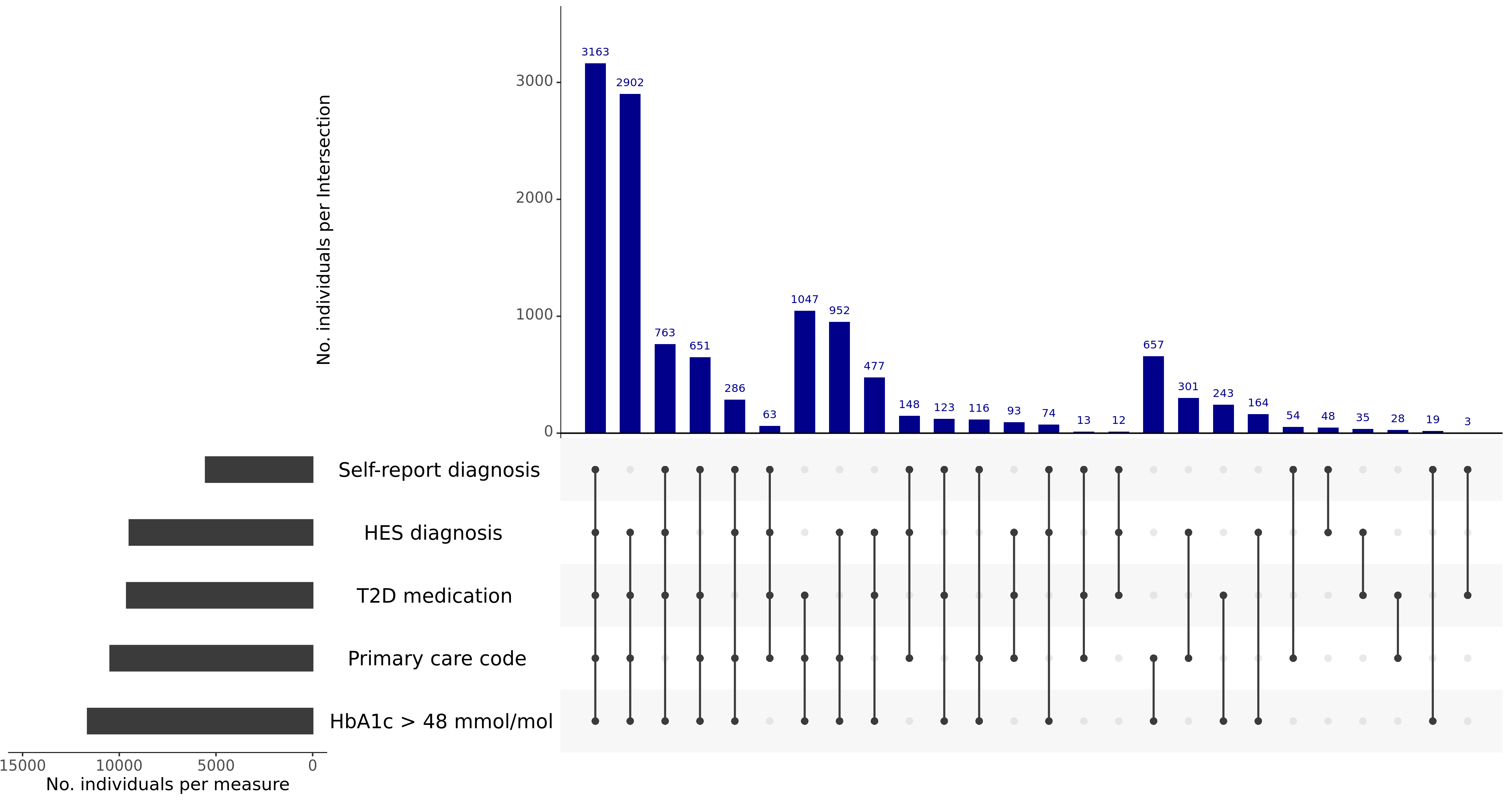
